# The evolving SARS-CoV-2 epidemic in Africa: Insights from rapidly expanding genomic surveillance

**DOI:** 10.1101/2022.04.17.22273906

**Authors:** Houriiyah Tegally, James E. San, Matthew Cotten, Bryan Tegomoh, Gerald Mboowa, Darren P. Martin, Cheryl Baxter, Monika Moir, Arnold Lambisia, Amadou Diallo, Daniel G. Amoako, Moussa M. Diagne, Abay Sisay, Abdel-Rahman N. Zekri, Abdelhamid Barakat, Abdou Salam Gueye, Abdoul K. Sangare, Abdoul-Salam Ouedraogo, Abdourahmane Sow, Abdualmoniem O. Musa, Abdul K. Sesay, Adamou Lagare, Adedotun-Sulaiman Kemi, Aden Elmi Abar, Adeniji A. Johnson, Adeola Fowotade, Adewumi M. Olubusuyi, Adeyemi O. Oluwapelumi, Adrienne A. Amuri, Agnes Juru, Ahmad Mabrouk Ramadan, Ahmed Kandeil, Ahmed Mostafa, Ahmed Rebai, Ahmed Sayed, Akano Kazeem, Aladje Balde, Alan Christoffels, Alexander J. Trotter, Allan Campbell, Alpha Kabinet Keita, Amadou Kone, Amal Bouzid, Amal Souissi, Ambrose Agweyu, Ana V. Gutierrez, Andrew J. Page, Anges Yadouleton, Anika Vinze, Anise N. Happi, Anissa Chouikha, Arash Iranzadeh, Arisha Maharaj, Armel Landry Batchi-Bouyou, Arshad Ismail, Augustina Sylverken, Augustine Goba, Ayoade Femi, Ayotunde Elijah Sijuwola, Azeddine Ibrahimi, Baba Marycelin, Babatunde Lawal Salako, Bamidele S. Oderinde, Bankole Bolajoko, Beatrice Dhaala, Belinda L. Herring, Benjamin Tsofa, Bernard Mvula, Berthe-Marie Njanpop-Lafourcade, Blessing T. Marondera, Bouh Abdi Khaireh, Bourema Kouriba, Bright Adu, Brigitte Pool, Bronwyn McInnis, Cara Brook, Carolyn Williamson, Catherine Anscombe, Catherine B. Pratt, Cathrine Scheepers, Chantal G. Akoua-Koffi, Charles N. Agoti, Cheikh Loucoubar, Chika Kingsley Onwuamah, Chikwe Ihekweazu, Christian Noël Malaka, Christophe Peyrefitte, Chukwuma Ewean Omoruyi, Clotaire Donatien Rafaï, Collins M. Morang’a, D. James Nokes, Daniel Bugembe Lule, Daniel J. Bridges, Daniel Mukadi-Bamuleka, Danny Park, David Baker, Deelan Doolabh, Deogratius Ssemwanga, Derek Tshiabuila, Diarra Bassirou, Dominic S.Y. Amuzu, Dominique Goedhals, Donald S. Grant, Donwilliams O. Omuoyo, Dorcas Maruapula, Dorcas Waruguru Wanjohi, Ebenezer Foster-Nyarko, Eddy K. Lusamaki, Edgar Simulundu, Edidah M. Ong’era, Edith N. Ngabana, Edward O. Abworo, Edward Otieno, Edwin Shumba, Edwine Barasa, El Bara Ahmed, Elizabeth Kampira, Elmostafa El Fahime, Emmanuel Lokilo, Enatha Mukantwari, Erameh Cyril, Eromon Philomena, Essia Belarbi, Etienne Simon-Loriere, Etilé A. Anoh, Fabian Leendertz, Fahn M. Taweh, Fares Wasfi, Fatma Abdelmoula, Faustinos T. Takawira, Fawzi Derrar, Fehintola V Ajogbasile, Florette Treurnicht, Folarin Onikepe, Francine Ntoumi, Francisca M. Muyembe, Francisco Ngiambudulu, Frank Edgard Zongo Ragomzingba, Fred Athanasius Dratibi, Fred-Akintunwa Iyanu, Gabriel K. Mbunsu, Gaetan Thilliez, Gemma L. Kay, George O. Akpede, Uwem E. George, Gert van Zyl, Gordon A. Awandare, Grit Schubert, Gugu P. Maphalala, Hafaliana C. Ranaivoson, Hajar Lemriss, Hannah E Omunakwe, Harris Onywera, Haruka Abe, Hela Karray, Hellen Nansumba, Henda Triki, Herve Albéric Adje Kadjo, Hesham Elgahzaly, Hlanai Gumbo, Hota mathieu, Hugo Kavunga-Membo, Ibtihel Smeti, Idowu B. Olawoye, Ifedayo Adetifa, Ikponmwosa Odia, Ilhem Boutiba-Ben Boubaker, Isaac Ssewanyana, Isatta Wurie, Iyaloo S Konstantinus, Jacqueline Wemboo Afiwa Halatoko, James Ayei, Janaki Sonoo, Jean Bernard Lekana-Douki, Jean-Claude C. Makangara, Jean-Jacques M. Tamfum, Jean-Michel Heraud, Jeffrey G. Shaffer, Jennifer Giandhari, Jennifer Musyoki, Jessica N. Uwanibe, Jinal N. Bhiman, Jiro Yasuda, Joana Morais, Joana Q. Mends, Jocelyn Kiconco, John Demby Sandi, John Huddleston, John Kofi Odoom, John M. Morobe, John O. Gyapong, John T. Kayiwa, Johnson C. Okolie, Joicymara Santos Xavier, Jones Gyamfi, Joseph Humphrey Kofi Bonney, Joseph Nyandwi, Josie Everatt, Jouali Farah, Joweria Nakaseegu, Joyce M. Ngoi, Joyce Namulondo, Judith U. Oguzie, Julia C. Andeko, Julius J. Lutwama, Justin O’Grady, Katherine J Siddle, Kathleen Victoir, Kayode T. Adeyemi, Kefentse A. Tumedi, Kevin Sanders Carvalho, Khadija Said Mohammed, Kunda G. Musonda, Kwabena O. Duedu, Lahcen Belyamani, Lamia Fki-Berrajah, Lavanya Singh, Leon Biscornet, Leonardo de Oliveira Martins, Lucious Chabuka, Luicer Olubayo, Lul Lojok Deng, Lynette Isabella Ochola-Oyier, Madisa Mine, Magalutcheemee Ramuth, Maha Mastouri, Mahmoud ElHefnawi, Maimouna Mbanne, Maitshwarelo I. Matsheka, Malebogo Kebabonye, Mamadou Diop, Mambu Momoh, Maria da Luz Lima Mendonça, Marietjie Venter, Marietou F Paye, Martin Faye, Martin M. Nyaga, Mathabo Mareka, Matoke-Muhia Damaris, Maureen W. Mburu, Maximillian Mpina, Mfoutou Mapanguy Claujens Chastel, Michael Owusu, Michael R. Wiley, Mirabeau Youtchou Tatfeng, Mitoha Ondo’o Ayekaba, Mohamed Abouelhoda, Mohamed Amine Beloufa, Mohamed G Seadawy, Mohamed K. Khalifa, Mohammed Koussai Dellagi, Mooko Marethabile Matobo, Mouhamed Kane, Mouna Ouadghiri, Mounerou Salou, Mphaphi B. Mbulawa, Mudashiru Femi Saibu, Mulenga Mwenda, Muluken Kaba, My V.T. Phan, Nabil Abid, Nadia Touil, Nadine Rujeni, Nalia Ismael, Ndeye Marieme Top, Ndongo Dia, Nédio Mabunda, Nei-yuan Hsiao, Nelson Boricó Silochi, Ngonda Saasa, Nicholas Bbosa, Nickson Murunga, Nicksy Gumede, Nicole Wolter, Nikita Sitharam, Nnaemeka Ndodo, Nnennaya A. Ajayi, Noël Tordo, Nokuzola Mbhele, Norosoa H Razanajatovo, Nosamiefan Iguosadolo, Nwando Mba, Ojide C. Kingsley, Okogbenin Sylvanus, Okokhere Peter, Oladiji Femi, Olumade Testimony, Olusola Akinola Ogunsanya, Oluwatosin Fakayode, Onwe E. Ogah, Ousmane Faye, Pamela Smith-Lawrence, Pascale Ondoa, Patrice Combe, Patricia Nabisubi, Patrick Semanda, Paul E. Oluniyi, Paulo Arnaldo, Peter Kojo Quashie, Philip Bejon, Philippe Dussart, Phillip A. Bester, Placide K. Mbala, Pontiano Kaleebu, Priscilla Abechi, Rabeh El-Shesheny, Rageema Joseph, Ramy Karam Aziz, René Ghislain Essomba, Reuben Ayivor-Djanie, Richard Njouom, Richard O. Phillips, Richmond Gorman, Robert A. Kingsley, Rosemary Audu, Rosina A.A. Carr, Saâd El Kabbaj, Saba Gargouri, Saber Masmoudi, Safietou Sankhe, Sahra Isse Mohamed, Salma Mhalla, Salome Hosch, Samar Kamal Kassim, Samar Metha, Sameh Trabelsi, Sanaâ Lemriss, Sara Hassan Agwa, Sarah Wambui Mwangi, Seydou Doumbia, Sheila Makiala-Mandanda, Sherihane Aryeetey, Shymaa S. Ahmed, Sidi Mohamed Ahmed, Siham Elhamoumi, Sikhulile Moyo, Silvia Lutucuta, Simani Gaseitsiwe, Simbirie Jalloh, Soafy Andriamandimby, Sobajo Oguntope, Solène Grayo, Sonia Lekana-Douki, Sophie Prosolek, Soumeya Ouangraoua, Stephanie van Wyk, Stephen F. Schaffner, Stephen Kanyerezi, Steve Ahuka-Mundeke, Steven Rudder, Sureshnee Pillay, Susan Nabadda, Sylvie Behillil, Sylvie L. Budiaki, Sylvie van der Werf, Tapfumanei Mashe, Tarik Aanniz, Thabo Mohale, Thanh Le-Viet, Thirumalaisamy P. Velavan, Tobias Schindler, Tongai Maponga, Trevor Bedford, Ugochukwu J. Anyaneji, Ugwu Chinedu, Upasana Ramphal, Vincent Enouf, Vishvanath Nene, Vivianne Gorova, Wael H. Roshdy, Wasim Abdul Karim, William K. Ampofo, Wolfgang Preiser, Wonderful T. Choga, Yahaya Ali Ahmed, Yajna Ramphal, Yaw Bediako, Yeshnee Naidoo, Yvan Butera, Zaydah R. de Laurent, Ahmed E.O. Ouma, Anne von Gottberg, George Githinji, Matshidiso Moeti, Oyewale Tomori, Pardis C. Sabeti, Amadou A. Sall, Samuel O. Oyola, Yenew K. Tebeje, Sofonias K. Tessema, Tulio de Oliveira, Christian Happi, Richard Lessells, John Nkengasong, Eduan Wilkinson

## Abstract

Investment in Africa over the past year with regards to SARS-CoV-2 genotyping has led to a massive increase in the number of sequences, exceeding 100,000 genomes generated to track the pandemic on the continent. Our results show an increase in the number of African countries able to sequence within their own borders, coupled with a decrease in sequencing turnaround time. Findings from this genomic surveillance underscores the heterogeneous nature of the pandemic but we observe repeated dissemination of SARS-CoV-2 variants within the continent. Sustained investment for genomic surveillance in Africa is needed as the virus continues to evolve, particularly in the low vaccination landscape. These investments are very crucial for preparedness and response for future pathogen outbreaks.

**One-Sentence Summary:** Expanding Africa SARS-CoV-2 sequencing capacity in a fast evolving pandemic.

## Introduction

What originally started as a small cluster of pneumonia cases in Wuhan, China over two years ago (*1*), quickly turned into a global pandemic. Coronavirus Disease 2019 (COVID-19) is the clinical manifestation of severe acute respiratory syndrome coronavirus 2 (SARS-CoV-2) infection; by March 2022 there had been over 437 million reported cases and over 5.9 million reported deaths (*2*). Though Africa accounts for the lowest number of reported cases and deaths thus far, with ∼11.3 million reported cases and 245 000 reported deaths as of February 2022, the continent has played an important role in shaping the scientific response to the pandemic with the implementation of genomic surveillance and the identification of two of the five variants of concerns (VOCs) (*3, 4*).

Since it emerged in 2019, SARS-CoV-2 has continued to evolve and adapt (*5*). This has led to the emergence of several viral lineages that carry mutations that confer some viral adaptive advantages that increase their odds of transmission and infection (*6, 7*), or counter the effect of neutralizing antibodies from vaccination (*8*) or previous infections (*9–11*). The World Health Organization (WHO) classifies certain viral lineages as VOCs or variants of interest (VOIs) based on the potential impact they may have on the pandemic, with VOCs regarded as the highest risk. To date, five VOCs have been classified by the WHO, two of which were first detected on the African continent (Beta and Omicron) (*3, 4, 12*), while two more (Alpha and Delta) (*12, 13*) have spread extensively on the continent in successive waves. The remaining VOC, Gamma (*14*), which originated in Brazil has had a limited influence in Africa with only four recorded sequenced cases.

For genomic surveillance to be useful for public health responses, sampling needs to be both spatially and temporally representative. In the case of SARS-CoV-2 in Africa, this means extending the geographic coverage of sequencing capacity to be able to capture the dynamic genomic epidemiology in as many locations as possible. In a meta-analysis of the first 10 000 SARS-CoV-2 sequences generated in 2020 from Africa (*15*) several blindspots were identified with regards to genomic surveillance on the continent. Since then, a lot of investment has been poured into building capacity for genomic surveillance in Africa coordinated mostly by the Africa Centers for Disease Control (Africa CDC) and the WHO AFRO, but also provided by several national and international partners, resulting in an additional 90,000 sequences shared over the past year. This makes the sequencing effort for SARS-CoV-2 a phenomenal milestone making it the fastest and most concentrated sequencing efforts for any human pathogen in the history of the continent and globally. In comparison, only 12,000 whole genome influenza sequences (*16*) and only ∼3,100 whole genome HIV sequences (*17*) have been shared publicly even though HIV has plagued the continent for decades.

Here we describe how the first 100,000 SARS-CoV-2 genomes from Africa have helped with understanding the epidemic on the continent, how this genomic surveillance in Africa expanded, and how we adapted our sequencing methods to deal with an evolving virus. We also highlight the impact that genomic sequencing in Africa has had on the global public health response, particularly through the identification and early analysis of new variants.

## Results

### Epidemic waves driven by variant dynamics and geography

Scaling up sequencing in Africa has provided a wealth of information on how the pandemic unfolded on the continent. The epidemic has largely been heterogeneous across the continent, but most countries have experienced multiple waves of infection (*18–29*), with significant local and regional diversity in the first and to a lesser extent the second waves, followed by successive sweeps of the continent with Delta and Omicron VOCs (**Figure 1A**). In all regions of the continent, different lineages and VOIs evolved and co-circulated with VOCs and in some cases, contributed considerably to epidemic waves.

**Figure 1:**
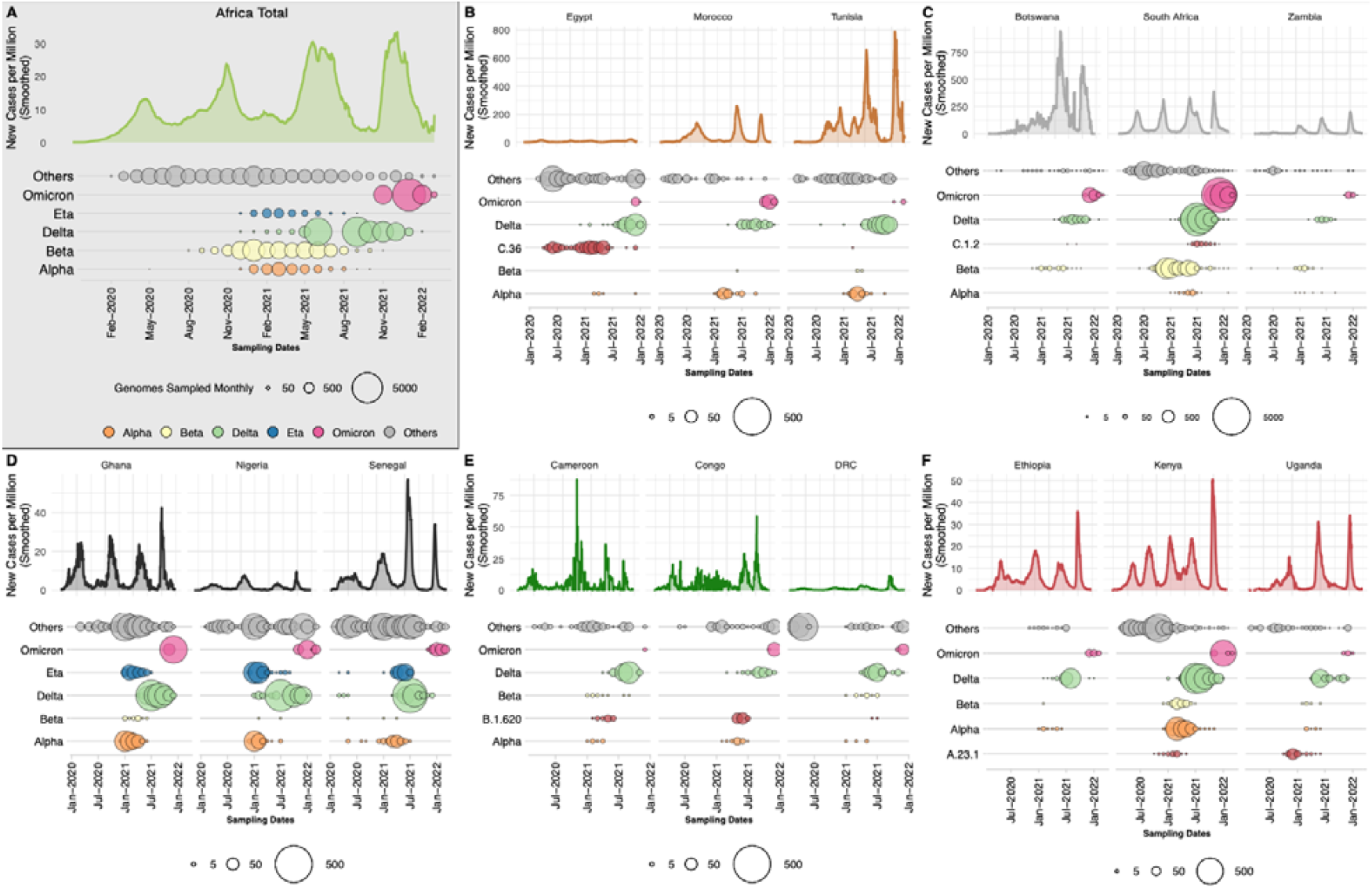
Epidemiological progression of the COVID-19 pandemic on the African continent. A) Total new case counts per million inhabitants in Africa along with the distribution of VOCs, the Eta VOI and other lineages through time (size of circles proportional to the number of genomes sampled per month for each category). (B-F) Breakdown of new cases per million and monthly sampling of VOCs, regional variant or lineage of interest and other lineages for three selected countries for North, Southern, West, Central and East Africa respectively. For each region, a different variant or lineage of interest is shown, relevant to that region (C.36, C.1.2, Eta, B.1.620 and A.23.1 respectively). Note that the y-axis scales differ between regional sub-plots.

In North Africa (**Figure 1B, Supp Figure S1A**), B.1 lineages and Alpha dominated the pandemic in the first and second wave of the pandemic and were replaced by Delta and Omicron in the third and fourth waves, respectively. Interestingly, the C.36 and C.36.3 sub-lineage dominated the epidemic in Egypt (∼40% of infections) before July 2021 when it was replaced by Delta (*30*). Similarly in Tunisia the first and second waves were associated with the B.1.160 lineage and were replaced by Delta during the country’s third wave of infections. In Southern Africa (**Figure 1C, Supp Figure S1C**), we see a similar pandemic profile with B.1 dominating the first wave, but instead of Alpha, Beta was responsible for the second wave, followed by Delta and Omicron. Another lineage that was flagged for close monitoring in the region was C.1.2, due to its mutational profile and predicted capacity for immune escape (*31*). However, the C.1.2 lineage did not cause many infections in the region as it was circulating at a time when Delta was dominant. In West Africa (**Figure 1D, Supp Figure S1B**), the B.1.525 lineage caused a large proportion of infections in the second and third waves where it shared the pandemic landscape with the Alpha variant. As with other regions on the continent, these variants were later replaced by the Delta and then Omicron VOCs in successive waves. In Central Africa (**Figure 1E, Supp Figure S1D**), the B.1.620 lineage caused most of the infections between January and June 2021 (*32*) before systematically being replaced by Delta and then Omicron. Lastly, in East Africa (**Figure 1F, Supp Figure S1E**) the A.23.1 lineage dominated the second wave of infections in Uganda (*33*) and much of East Africa. In all of these regions, minor lineages such as B.1.525, C.36 and A.23.1 were eventually replaced by VOCs that emerged

Finally, we directly compared the official recorded cases in Africa with the ongoing SARS-CoV-2 genomic surveillance (GISAID date of access 2022-02-15) for a crude estimation of variants’ contribution to cases, with the obvious caveats that testing and reporting practices vary widely across the continent and genomic surveillance volumes have varied throughout the pandemic. We observe that Delta was responsible for an epidemic wave between May and October 2021 (**Figure 1A**) and had the greatest impact on the continent with almost 38.5% of overall infections in Africa possibly attributed to it. The second most impactful variant was Beta, responsible for an epidemic wave at the end of 2020 and beginning of 2021 (**Figure 1A**), with 15.7% of infections overall attributed to it. Notably, Alpha, despite being predominant in other parts of the world at the beginning of 2021, had only minimal significance in Africa, accounting for just 4.7% of infections.

At the time of writing, the Omicron wave was still unfolding globally and in Africa and its full impact may turn out to be much bigger. However, due to increased population immunity, from SARS-CoV-2 infection and vaccination, the impact of Omicron on mortality has been less than with the other VOCs, as can be observed by the relatively low death rates in South Africa during the Omicron wave (*34*). The findings from mapping epidemiological numbers onto genomic surveillance data are reliable given that the sampling of genomes across Africa appears to scale proportionally to the size and timing of epidemic waves (**Supp Figure S2**).

Regional (**Supplementary Table S1**) and country (**Supplementary Table S2**) specific Nextstrain builds for Africa also clearly support the changing nature of the pandemic over time with B.1-like viruses circulating during the first wave, and were replaced by either the Alpha or Beta variant in the second wave, the Delta variant in the third wave and Omicron in the fourth wave.

### Phylogenetic insights into the rise and spread of Variants of Concern in Africa

During the first wave of infections in 2020 in Africa, as was the case globally, the majority of corresponding genomes were classified as PANGO B.1 (n=2 456) or B.1.1 viruses (n=1 329). Towards the end of 2020, more distinct viral lineages started to appear. The most important of which that impacted the African continent are: B.1.525 (n=797), B.1.1.318 (n=398) (*35*), B.1.1.418 (n=395), A.23.1 (n=358) ((*15, 29, 31, 33*)), C.1 (n=446) (*29*), C.1.2 (n=300) (*31*), C.36 (n=305) (*30, 36*), B.1.1.54 (n=287) ((*15, 29, 31, 33*)), B.1.416 (n=272), B.1.177 (n=203), B.1.620 (n=138), and B.1.160 (n=61), (*32*) (**Supp Fig S3A,B**). From our discrete state phylogeographic inference we infer at least 2,151 (95% CI: 2,132 - 2,170) viral introductions of non-VOC lineages into African countries over the course of the pandemic. While there was an initial dominance of introductions from Europe, 62% of the overall viral introductions (n=1 333; 95% CI: 1,319 - 1,347) were attributed to another African country (**Supp Fig S3C,D**). Of the 38% foreign viral introductions (n=818; 95% CI: 805 - 831) the bulk were attributed to the United Kingdom UK (∼255), US (∼179), and EU countries (∼176). Conversely, 930 (95% CI: 919 - 941) viral exports from Africa to countries outside the continent were observed with the bulk attributed to Europe (∼432), North America (∼213) and Asia (∼195).

As with the unfolding of events globally, VOCs became increasingly important to the epidemic in Africa towards the end of 2020. The Alpha, Beta, Delta and Omicron variants demonstrate many similarities as well as differences in the way they spread on the continent. For all these VOCs, we observe large regional monophyletic transmission clusters in each of their phylogenetic reconstructions in Africa (**Supp Fig S4**). This suggests an important extent of continental dissemination within Africa. While the Alpha and Beta variants were epidemiologically important in distinct regions of the continent, the Delta and Omicron variants sequentially dominated the majority of infections on the entire continent shortly after their emergence (**Figure 2A)**.

**Figure 2:**
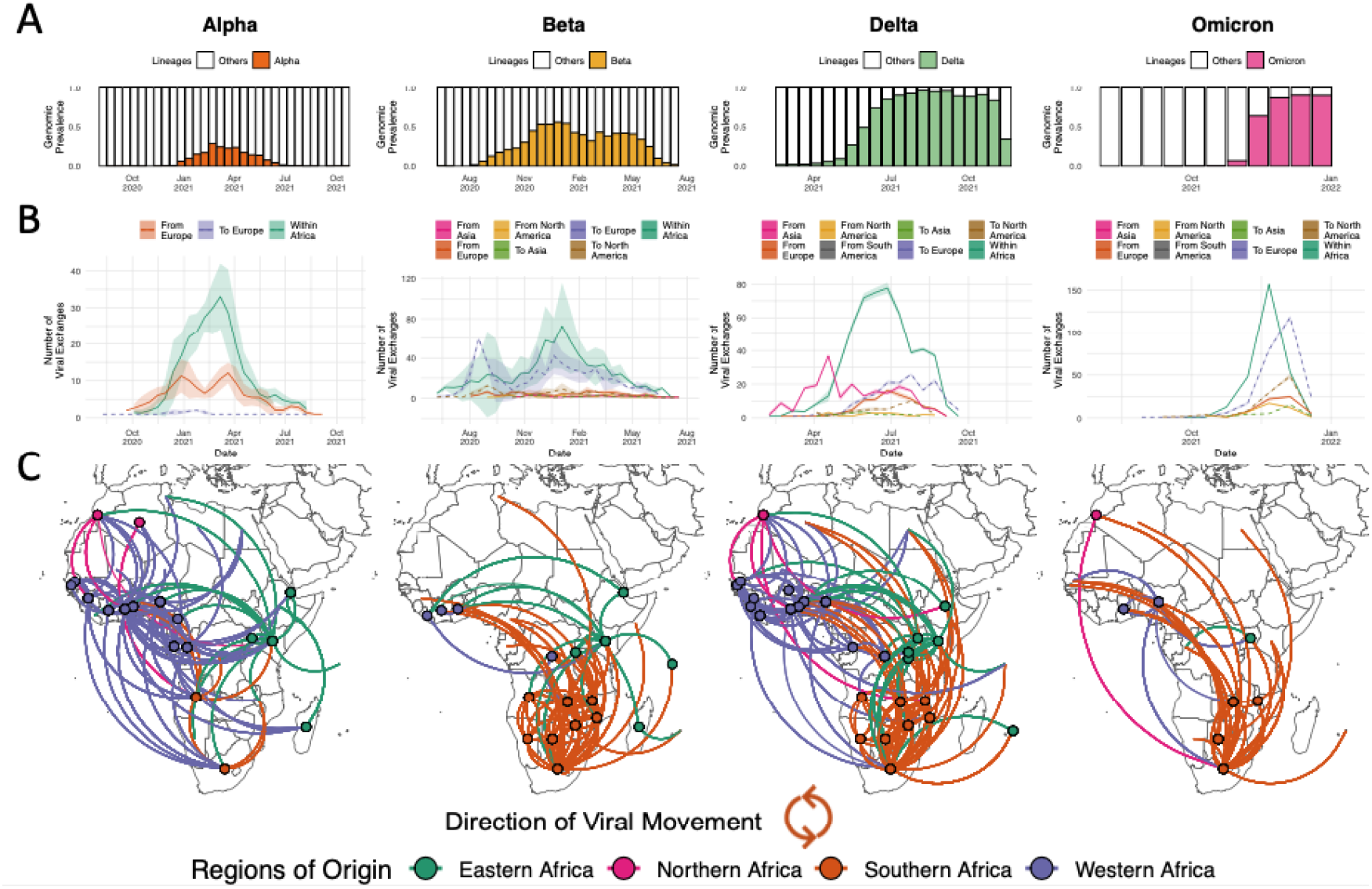
Inferred viral dissemination patterns of VOCs within Africa. A) Genomic prevalence of VOCs Alpha, Beta, Delta and Omicron in Africa over time. B) Inferred viral exchange patterns to, from and within the Africa continent for the four VOCs. Introductions and viral transitions within Africa are shown in solid lines and exports from Africa are shown in dotted lines and these are coloured by continent. The shaded areas around the lines represent uncertainty of this analysis from ten replicates (+/- s.d.). Note that the y-axis scales differ between subplots. C) Dissemination patterns of the VOCs within Africa, from inferred ancestral state reconstructions, annotated and coloured by region in Africa. The countries of origin of viral exchange routes are also shown with dots and the curves go from country of origin to destination country in an anti-clockwise direction.

The Alpha variant was first identified in December 2020 in the UK and has since spread globally. In Africa, Alpha has been detected in 43 countries with evidence of community transmission, based on phylogenetic clustering, in many countries including Ghana, Nigeria, Kenya, Gabon and Angola. Even in southern Africa where the Beta variant was dominating the pandemic landscape at the time, Alpha still managed to cause significant community transmission (**Supp Fig S4**). Discrete state maximum likelihood reconstruction inferred at least 155 introductions (95% CI: 152 - 159) into Africa with the bulk of imports attributed to the UK (>97%) (**Figure 2B**). However, >66% of imports into any particular African country were actually attributed to another African nation, suggesting substantial dispersal of the Alpha variant within the continent, mostly originating from West African countries (∼71% of viral transfers within Africa) (**Figure 2C**). This is in spite of initial importations of the Alpha variant from Europe into all regions of the continent (**Supp Fig S5B**).

The second VOC, Beta, was identified in December 2020 in South Africa (*4*). However, sampling and molecular clock analyses suggest that the variant originated around September 2020 in South Africa (**Supp Fig S4**). At the end of 2020 and beginning of 2021, Beta was driving a second wave of infection in South Africa and quickly spread to other countries within the region. The concurrent introductions and spread of Alpha and other variants (Eta, A.23.1) in other regions of the continent substantially reduced the Beta variant’s initial growth, limiting its spread to largely Southern Africa, and to a lesser extent the East Africa regions. Beta spread to at least 114 countries globally, including 37 countries and territories in Africa. Of the 900 (95% CI: 895 - 905) inferred introductions of the Beta variant into African countries, only 108 (95% CI: 105 - 111) were attributed to sources outside the continent (**Supp Fig5C**), while nearly half of introductions were attributed to South Africa (51.3%) (**Figure 2C**). This is in line with expectations as the variant originated in South Africa and the dominance of Alpha in most other parts of the world, which may have limited the international spread of the Beta variant (**Figure 2B**). Beyond southern Africa, most of the introductions back into the African continent were attributed to France and other EU countries into the French overseas territories, Mayotte and Reunion, and other Francophone African countries.

The fourth VOC was Delta (*13*), which rose to prominence in April 2021 in India, where it fueled an explosive second wave. Since its emergence, Delta was detected in >170 countries, including 37 African countries and territories (**Supp Fig S4**). Our analysis infers at least 399 (95% CI: 393 - 405) introductions of the Delta variant into Africa, with the bulk attributed to India (∼52%), mainland Europe (∼25%), the UK (∼14%), and the US (∼6%). However, as with the Alpha variant, while all regions of the continent saw a large number of introductions from Asia (**Supp Fig S5D**), the overall majority of Delta introductions into any African country were in fact from another African country (mean 67%; 95% CI: 65 - 69%) (**Figure 1B**), suggesting evidence of rapid spread within and between regions. We infer that viral dissemination of Delta within Africa was not restricted to or dominated by any particular region unlike Alpha and Beta, but rather spread across the entire continent (**Figure 1C**). Following introductions from Asia in the middle of 2021, Delta rapidly replaced the other circulating variants. For example, in southern African countries, the Delta variant rapidly displaced Beta and by June-2021 was circulating at very high (>90%) frequencies (*37*).

The latest VOC, Omicron, was identified and characterized in November 2021, in southern Africa (*3*). At the time of writing, the variant has been detected in >150 countries including 36 African countries and one overseas territory (**Supp Fig S4**). Our discrete ancestral state reconstruction infers at least 397 (95% CI: 391 - 403) introductions of the Omicron variant into various African countries, of which only 121 (95% CI: 115 - 127) were attributed to sources outside of the continent (**Figure 1B**). Similar to Beta, viral dissemination of Omicron within the continent mostly originated from southern Africa (**Figure 1C**). Outside introductions of the Omicron back into the African continent were dominated by the UK (∼47%), USA (∼33%), Australia and New Zealand (∼9%). These reintroductions of the Omicron variant back into Africa were primarily directed to countries outside of the southern African region (e.g. Nigeria, Ghana or Kenya) where the variant was first identified (**Supp Fig 5D**). As expected, we also infer a considerable proportion of viral exportation events of Omicron from Africa to Europe (**Figure 1B**).

### Optimizing surveillance coverage in Africa

By mapping and comparing the locations of specimen sampling laboratories to the sequencing laboratories, a number of aspects regarding the expansion of genomic surveillance on the continent became clear. First, even though several countries in Africa started sequencing SARS-CoV-2 in the first months of the pandemic, local sequencing capacity was initially limited. However, local sequencing capabilities slowly expanded over time, particularly after the emergence of VOCs (**Figure 3A**). The fact that almost half of all SARS-CoV-2 sequencing in Africa was performed using the Oxford Nanopore technology (ONT), which is relatively low-cost compared to other sequencing technologies and better adapted to modest laboratory infrastructures, illustrates one component of how this rapid scale-up of local sequencing was achieved (**Supp Fig S6**). Yet, to rely only on local sequencing would have thwarted the continent’s chance at a reliable genomic surveillance program. At the time of writing, there were 52/55 countries in Africa with SARS-CoV-2 genomes deposited in GISAID, however, there were still 16 countries with no reported local sequencing capacity (**Figure 3A**) and undoubtedly many with limited capacity for the demands during pandemic waves.

**Figure 3:**
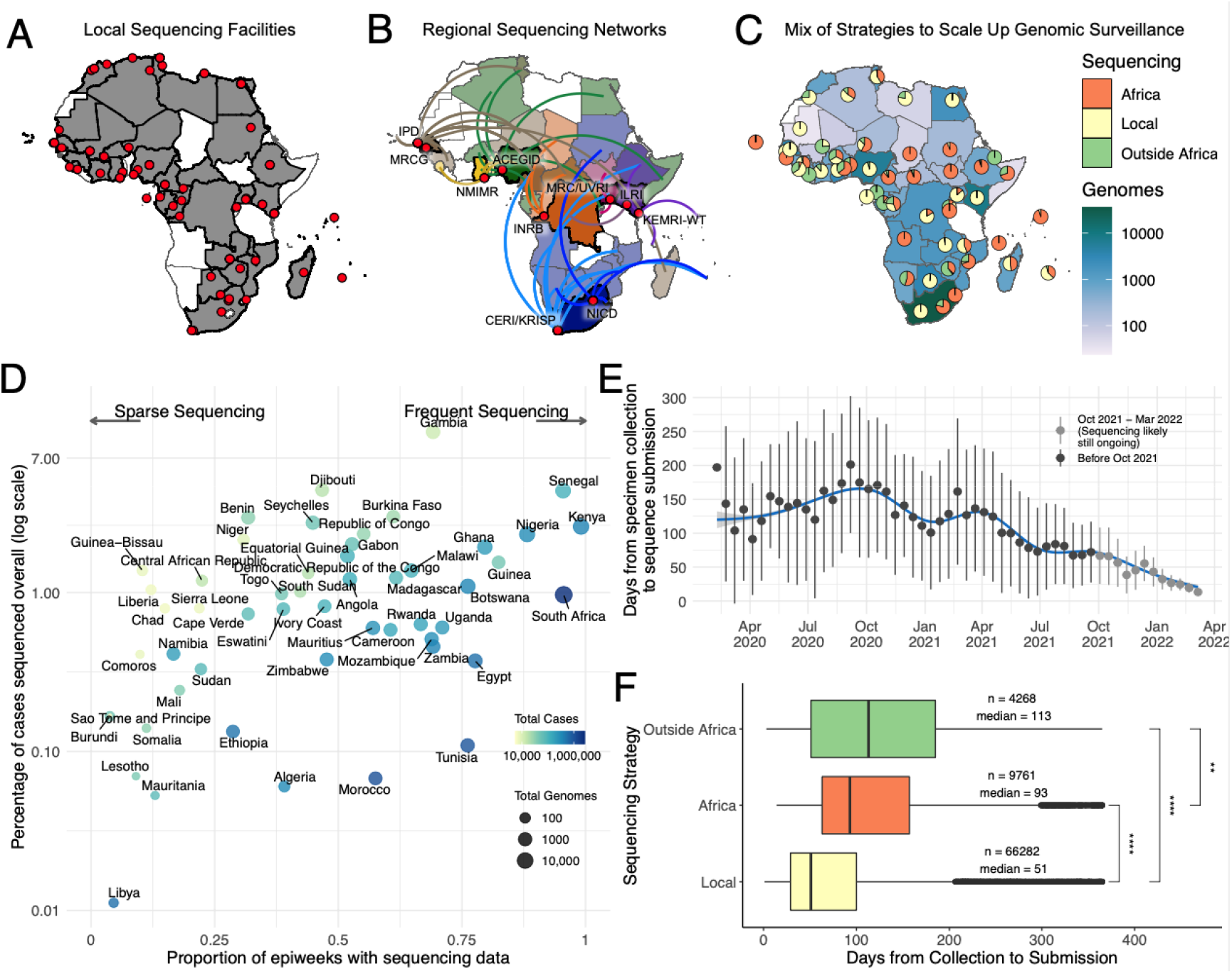
Sequencing strategies and outputs in Africa. A) Geographical representation of all countries (shaded in gray) and institutions (red dots) in Africa with their own on-site sequencing facilities. B) Key regional sequencing hubs and networks in Africa showing countries (shaded in bright colors) and institutions (red dots) that have sequenced for other countries (shaded in corresponding light colors and linking curves) on the continent. CERI: Centre for Epidemic Response and Innovation; KRISP: KwaZulu-Natal Research Innovation and Sequencing Platform; NICD: National Institute for Communicable Diseases; KEMRI-WT: Kenya Medical Research Institute - Wellcome Trust; ILRI: International Livestock Research Institute; MRC/UVRI: Medical Research Council/Uganda Virus Research Institute; INRB: Institut National de Recherche Biomédicale; ACEGID: African Centre of Excellence for Genomics of Infectious Diseases; NMIMR: Noguchi Memorial Institute for Medical Research; MRCG: Medical Research Council Unit - The Gambia; IPD: Institut Pasteur de Dakar C) Geographical representation of the total number of SARS-CoV-2 whole genomes produced over the course of the pandemic in each country as well as the proportion of those sequences that were produced locally, regionally or abroad. D) Correlation of the proportion of COVID-19 positive cases that have been sequenced and the corresponding number of epidemiological weeks since the start of the pandemic that are represented with genomes for each African country. The color of each circle represents the number of cases and its size the number of genomes. E) Progression of sequencing turnaround time (calculated at the difference between the date of sampling and date of sequencing submission of each sample) for the whole continent over the course of the pandemic. The dot and error bars for each group denote the mean and range for two s.d., respectively F) Comparison of sequencing turn-around times (lag times from sample collection to submission) for the three strategies of sequencing in Africa, showing a significant difference in the means (p-value<0.0001). The box and whisker plot denote the lower quartile, median and upper quartile (box), the minimum maximum values (whisker), and outliers (black dots)

To tackle this, regional sequencing hubs were established aimed at maximizing resources available in a few countries to assist in genomic surveillance across the continent. This sequencing is done either as the sole source of viral genomes for those countries (e.g. Angola, South Sudan and Namibia) or concurrently with local efforts to increase capacity during resurgences (**Figure 3B**). Sequencing is further supplemented by a number of countries utilizing facilities outside of Africa. Ultimately, a mix of strategies from local sequencing, collaborative resource sharing among African countries and sequencing with academic collaborators outside the continent helped close surveillance blindspots (**Figure 3C**). Countries in sub-Saharan Africa, particularly in Southern and East Africa, most benefited from the regional sequencing networks, while countries in West and North Africa often partnered with collaborators outside of Africa.

The success of pathogen genomic surveillance programs relies on how representative it is of the epidemic under investigation. For SARS-CoV-2, this is often measured in terms of the percentage of reported cases sequenced and the frequency of sampling. African countries were positioned across a range of different combinations of overall proportion and frequency of genomic sampling (**Figure 3D**). While the ultimate goal would be to optimize both of these parameters, a lower proportion of sampling can also be useful if frequency of sampling is maintained as high as possible. For instance, South Africa and Nigeria, who have both sequenced ∼1% of cases overall, can be considered to have successful genomic surveillance programs on the basis that sampling is representative over time, and has enabled the timely detection of variants (Beta, Eta, Omicron).

Additionally, for genomic surveillance to be most useful for rapid public health response during a pandemic, sequencing would ideally be done in real-time or in a framework as close as possible to that. We show that sequencing turn-around time in Africa improved (**Figure 3E**), decreasing from a mean of 171 days between October to December 2020 to a mean of 32 days over the same period a year later, although this does come with several caveats. First, we measure sequencing turn-around time in the most accessible manner, which is by comparing the date of sampling of a specimen to the date its sequence is deposited in GISAID. Overall, genomic data potentially informs the public health response more rapidly than reflected here, particularly when it comes to local outbreak investigations or variant detection. This analysis is also confounded by various factors such as country-to-country variation in these trends (**Supp Figure S7**), delays in data sharing, and potential retrospective sequencing, particularly by countries joining sequencing efforts at later stages of the pandemic. The most critical caveat is the fact that sequencing from the most recently collected samples (e.g. over the last four months) may still be ongoing. The shortening duration between sampling and genomic data sharing is nevertheless a positive takeaway, given that this data also feeds into continental and global genomic monitoring networks. Overall, the continental average delay from specimen collection to sequencing submission is 87 days with 10 countries having an average turnaround time of less than 60 days and Botswana of less than 30 days (**Supp Figure S8**). As expected, we found that the route taken to sequencing matters to the speed of data generation. Local sequencing has significantly faster sequencing turn-around times of the three frameworks we investigated (median of 51 days), followed by sequencing within regional sequencing networks in Africa (median of 93 days) and finally outsourced sequencing to countries outside Africa (median of 113 days) (**Figure 3F**). These findings support the investments in local genomic surveillance, to generate timely data for local and regional decision making.

With the increase in sequencing capacity on the continent, a decrease in the time taken to detect new variants was observed. For example, the Beta variant was identified in December 2020 in South Africa (*4*), but sampling and molecular clock analyses suggest the variant originated in September 2020. This three-month lag in detection means that a new variant, like Beta, has ample time to spread over a large geographic region prior to its detection. However, by the end of 2021, the time to detect a new variant was substantially improved. Phylogenetic and molecular clock analyses suggest that the Omicron variant originated around 9 October 2021 (95% Highest posterior density or HPD: 30 September - 20 October 2021) and the variant was described on 23rd November 2021 (*3*). Thus, Omicron was detected within ∼5 weeks from origin compared to the Beta variant (∼16 weeks) and the Alpha variant, detected in the UK (∼10 weeks). More importantly, the time between sequence deposition to the WHO declaring the new variant a VOC was substantially shortened to 72 hours for the Omicron variant.

### Optimizing and developing sequencing protocols in Africa

Yet, the rapidity of SARS-CoV-2 evolution has complicated sequencing efforts. Common methods of RNA sequencing include reverse transcription followed by double stranded DNA amplification using sequence-specific primer sets (*38*). Ongoing SARS-CoV-2 evolution has necessitated the continual evaluation and updating of these primer sets to ensure their sustained utility during genomic surveillance efforts.

In addition to increasing the volume of genomic surveillance on the continent, it is critical to also maintain sequencing quality. For the sequencing of fast-evolving pathogens like SARS-CoV-2, viral evolution is an important factor to consider in the process of designing primers. Here, we examined the current set of genomes to determine aspects of the sequencing that might be improved upon in the future. Many of the primer sets used were designed using viral sequences at the start of the pandemic and may require updating to keep pace with evolution. Indeed, the ARTIC primer sets are currently in version 4.1 (*39*). The Entebbe primer set was designed mid-2020 way into the first year of the epidemic and used an algorithm and design that accommodates evolution (*40*). The effects of viral evolution on sequencing patterns can be seen with low median N values for the first 12 months of the epidemic with an increase from October 2020 (**Figure 4A**). Additional challenges appear (indicated by increasing median N values) as the virus further evolved to adapt to humans and avoid immune responses (**Figure 4A**). Examining the role of sequencing technology, it appears that the two major technologies used (Illumina and ONT) have similar gap profiles while Ion Torrent, MGI and Sanger show reduced N content (**Figure 4B**). Likely factors for this pattern are the primers used in sequencings, with primer choice playing a key role in the presence of gaps (**Figure 4C**). The position of gaps shows an enrichment in the genome regions after position 19 000 with frequent gaps disrupting the spike coding region (**Figure 4D**).

**Figure 4:**
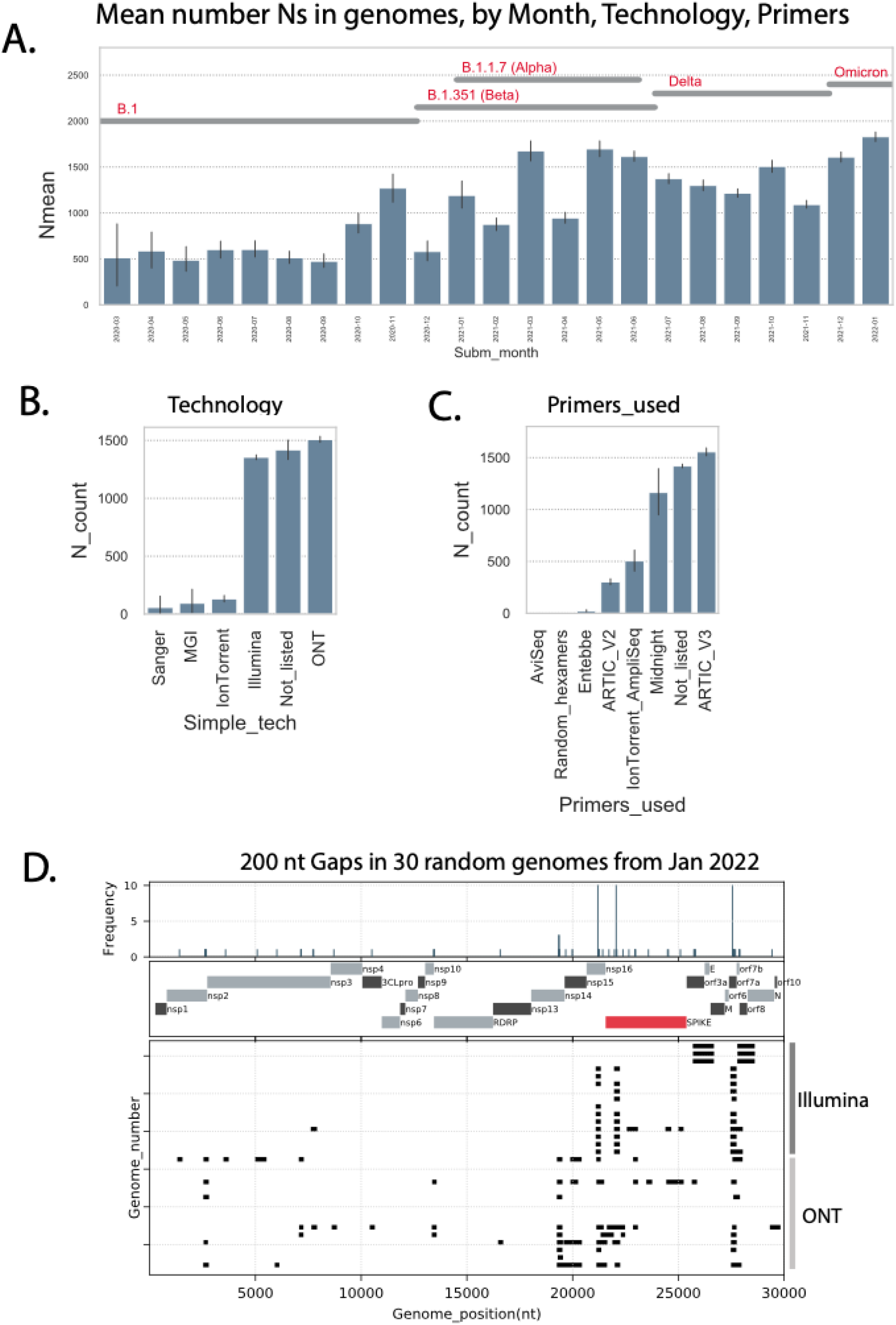
Genome gap analysis. A) Shows the mean N count per genome by month of submission to GISAID. The dates for the detection of important SARS-CoV-2 lineages are indicated at the top of the figure. B) Illustrates the mean N count per genome stratified by sequencing technology. C) Shows the mean N count per genome stratified by the sequencing primers sets used. For panels A to C, error bars indicate 95% confidence intervals. D) Shows the genome locations of 200nt gaps (strings of 200N) in 30 random African genomes from January 2022 (15 generated using Illumina methods, 15 with ONT). The upper histogram displays frequency of 200 base pair gaps (by start nt of gap), the middle graph shows SARS-CoV-2 genome features, bottom graph shows the positions of 200 nt gaps (black bars) with each vertical line a single genome.

## Conclusions

The investment in genomic surveillance in Africa has started bearing fruit. It should be intensified as the virus continues to evolve, requiring the adaptation of methodologies locally to keep pace with the emergence of variants. The continued development of sequencing protocols in Africa is of crucial importance (*40–42*) given the number of variants and lineages that emerged in, and were introduced to, the continent. In Northern Africa, the SARS-CoV-2 pandemic was caused by waves of infections that were similar to those seen in Europe (first wave = B.1 descendents, second wave = Alpha, third wave = Delta and firth wave = Omicron), in southern Africa the pattern was similar but with a Beta wave instead of an Alpha one. In East Africa, the pandemic was more complex, involving both Alpha and Beta as well as its own lineage A.23.1 before the arrival of Delta and Omicron. Central Africa experienced epidemic patterns sometimes mirroring East Africa and other times southern Africa. In West Africa Eta made a significant contribution to both a second wave (together with alpha) and a third wave (together with Delta).

In spite of the recent successful expansion of genomics surveillance in Africa, additional work remains necessary. Even with the Africa CDC - Africa PGI’s investment, there are still 16 countries with no sequencing capacity within their own borders. These countries’ only option is to send samples to continental sequencing hubs or to centers outside of the continent, which increases the turn-around times and limits the utility of genomic surveillance for public health decision making. Secondly, not all countries are willing to share data openly in a timely fashion for fear of being subject to travel bans or restrictions which could bring substantial economic harm. Such hesitancy has obvious potential ramifications for the future of genomic surveillance on the continent.

Sampling remains heterogeneous across the continent with high relative sampling frequencies in more developed countries such as South Africa, while consistent temporally and geographically representative sampling remains elusive in most of the countries on the continent. This heterogeneous sampling creates challenges for accurate epidemic reconstruction through phylogenetic means, in particular when inferring viral dispersal patterns as uneven sampling can bias results. Unfortunately, uneven sampling will continue and can only be addressed through sensitivity analyses such as the downsampling strategies we previously performed (*15*). There also exists the limitation that our genomic epidemiology understanding is influenced by the low level of testing in most if not all countries in Africa. Yet, this limitation may not be so problematic as even when countries have much higher coverage of cases/sequences (Denmark, Australia, UK), the findings may not differ substantially especially when surveillance is consistent and regular like we demonstrate happened in several African countries.

Africa needs to continue expanding genomic sequencing technologies on the continent. This holds true not just for SARS-CoV-2 but for other emerging or re-emerging pathogens on the continent. For example, WHO announced in February 2022 the re-emergence of wild polio in Africa, while sporadic influenza H1N1 and Ebola outbreaks continue to emerge on the continent. The Africa CDC has estimated that over 200 pathogen outbreaks are reported across the continent every year. Beyond the current pandemic, continued investment in these areas could serve the public health of the continent well into the 21st century.

## Data Availability

All of the SARS-CoV-2 whole genome sequences that were analyzed in the present study are all publicly available on the GISAID sequence database. A full list of the African sequences as well as global references are presented and acknowledged in Supplementary Table S3 and in our data repository, which will be made public at final publication. The github repo also contains all of the alignments, raw and time scaled ML tree topologies, data analysis and visualization scripts used here which will allow for the independent reproduction of results.

## Acknowledgments

First and foremost, we acknowledge authors in institutions in Africa and beyond who have made invaluable contributions towards specimen collection and sequencing to produce and share, via GISAID, SARS-CoV-2 genomic data from Africa. A full list of these supplementary authors can be found in the Supplementary Information section. We also acknowledge the authors from the originating and submitting laboratories worldwide, who generated and shared SARS-CoV-2 sequence data, via GISAID, from other regions in the world, which was used to contextualize the African genomic data (Supplementary Table S3).

## Author contribution

Conceptualization: HT, CB, SKT, TdO, RL, EW

Methodology: HT, TdO, RL, EW, JES, MC, BT, GM, DPM, AL, GG

Genomic Data Generation: HT, TdO, RL, EW, JES, MC, BT, GM, DPM, AL, GG, SKT, AD, DGA, MMD, AS, ANZ, AB, ASG, AKS, AO, AS, AOM, AKS, AL, AK, AEA, AAJ, AF, AMO, AOO, AAA, AJ, AMR, AK, AM, AR, AS, AK, AB, AC, AJT, AC, AKK, AK, AB, AS, AA, AVG, AJP, AY, AV, ANH, AC, AI, AM, ALB, AI, AS, AG, AF, AES, AI, BM, BLS, BSO, BB, BD, BLH, BT, BM, BN, BTM, BAK, BK, BA, BP, BM, CB, CW, CA, CBP, CS, CGA, CNA, CL, CKO, CI, CNM, CP, CEO, CDR, CMM, DJN, DBL, DJB, DM, DP, DB, DD, DS, DT, DB, DSA, DG, DSG, DOO, DM, DWW, EF, EKL, ES, EMO, ENN, EOA, EO, ES, EB, EBA, EEF, EL, EM, EC, EP, EB, ES, EAA, FL, FMT, FW, FA, FTT, FD, FVA, FR, FO, FN, FMM, FN, FEZR, FAD, FI, GKM, GT, GLK, GOA, GUE, GvZ, GAA, GS, GPM, HCR, HL, HEO, HO, HA, HK, HN, HT, HAAK, HE, HG, Hm, HK, IS, IBO, IA, IO, IBB, IS, IW, ISK, JWAH, JA, JS, JBL, JCM, JMT, JH, JGS, JG, JM, JNU, JNB, JY, JM, JQM, JK, JDS, JH, JKO, JMM, JOG, JTK, JCO, JSX, JG, JHKB, JN, JE, JF, JN, JMN, JN, JUO, JCA, JJL, JO, KJS, KV, KTA, KAT, KSC, KSM, KGM, KOD, LB, LF, LS, LB, LdOM, LC, LO, LLD, LIO, MM, MR, MM, ME, MM, MIM, MK, MD, MM, MdLLM, MV, MFP, MF, MMN, MM, MD, MWM, MM, MMCC, MO, MRW, MYT, MOA, MA, MAB, MGS, MKK, MKD, MM, MMM, MK, MO, MS, MBM, MFS, MM, MVP, NA, NT, NR, NI, NMT, ND, NM, NH, NBS, NS, NB, NM, NG, NW, NS, NN, NAA, NT, NM, NHR, NI, NM, OCK, OS, OP, OF, OT, OAO, OF, OEO, OF, PS, PO, PC, PN, PS, PEO, PA, PKQ, PB, PD, PAB, PKM, PK, PA, RE, RJ, RKA, RGE, RA, RN, ROP, RG, RAK, RA, RAC, SEK, SG, SM, SS, SIM, SM, SH, SKK, SM, ST, SL, SHA, SWM, SD, SM, SA, SSA, SMA, SE, SM, SL, SG, SJ, SA, SO, SG, SL, SP, SO, SvW, SFS, SK, SA, SR, SP, SN, SB, SLB, SvdW, TM, TA, TM, TL, TPV, TS, TM, TB, UJA, UC, UR, VE, VN, VG, WHR, WAK, WKA, WP, WTC, YAA, YR, YB, YN, YB, ZRdL, AEO, AvG, MM, OT, PCS, AAS, SOO, YKT, CH, JN

Data Analysis: HT, TdO, RL, EW, JES, MC, BT, GM, DPM, AL, GG, MM, SvW

Funding acquisition: TdO, GG, SKT, AEO, AvG, MM, OT, AAS, SOO, YKT, CH

Project administration: TdO, GG, SKT, AEO, AvG, MM, OT, AAS, SOO, YKT, CH, RL, EW, PCS, JN, GM, AD, DGA, MMD, AC, DWW, HO, SWM

Supervision: TdO, GG, SKT, AEO, AvG, MM, OT, AAS, SOO, YKT, CH, RL, EW, PCS, JN

Writing – original draft: TdO, SKT, RL, EW, GM, HT, JES, MC, DPM, CB

Writing – review & editing: TdO, SKT, RL, EW, GM, HT, JES, MC, DPM, CB, GG, AEO, AvG, MM, OT, AAS, SOO, YKT, CH, PCS, JN, AD, DGA, MMD, AC, DWW, HO, SWM, BT, AL, AS, ANZ, AB, ASG, AKS, AO, AS, AOM, AKS, AL, AK, AEA, AAJ, AF, AMO, AOO, AAA, AJ, AMR, AK, AM, AR, AS, AK, AB, AJT, AC, AKK, AK, AB, AS, AA, AVG, AJP, AY, AV, ANH, AC, AI, AM, ALB, AI, AS, AG, AF, AES, AI, BM, BLS, BSO, BB, BD, BLH, BT, BM, BN, BTM, BAK, BK, BA, BP, BM, CB, CW, CA, CBP, CS, CGA, CNA, CL, CKO, CI, CNM, CP, CEO, CDR, CMM, DJN, DBL, DJB, DM, DP, DB, DD, DS, DT, DB, DSA, DG, DSG, DOO, DM, EF, EKL, ES, EMO, ENN, EOA, EO, ES, EB, EBA, EEF, EL, EM, EC, EP, EB, ES, EAA, FL, FMT, FW, FA, FTT, FD, FVA, FR, FO, FN, FMM, FN, FEZR, FAD, FI, GKM, GT, GLK, GOA, GUE, GvZ, GAA, GS, GPM, HCR, HL, HEO, HA, HK, HN, HT, HAAK, HE, HG, Hm, HK, IS, IBO, IA, IO, IBB, IS, IW, ISK, JWAH, JA, JS, JBL, JCM, JMT, JH, JGS, JG, JM, JNU, JNB, JY, JM, JQM, JK, JDS, JH, JKO, JMM, JOG, JTK, JCO, JSX, JG, JHKB, JN, JE, JF, JN, JMN, JN, JUO, JCA, JJL, JO, KJS, KV, KTA, KAT, KSC, KSM, KGM, KOD, LB, LF, LS, LB, LdOM, LC, LO, LLD, LIO, MM, MR, MM, ME, MM, MIM, MK, MD, MM, MdLLM, MV, MFP, MF, MMN, MM, MD, MWM, MM, MMCC, MO, MRW, MYT, MOA, MA, MAB, MGS, MKK, MKD, MM, MMM, MK, MO, MS, MBM, MFS, MM, MVP, NA, NT, NR, NI, NMT, ND, NM, NH, NBS, NS, NB, NM, NG, NW, NS, NN, NAA, NT, NM, NHR, NI, NM, OCK, OS, OP, OF, OT, OAO, OF, OEO, OF, PS, PO, PC, PN, PS, PEO, PA, PKQ, PB, PD, PAB, PKM, PK, PA, RE, RJ, RKA, RGE, RA, RN, ROP, RG, RAK, RA, RAC, SEK, SG, SM, SS, SIM, SM, SH, SKK, SM, ST, SL, SHA, SD, SM, SA, SSA, SMA, SE, SM, SL, SG, SJ, SA, SO, SG, SL, SP, SO, SvW, SFS, SK, SA, SR, SP, SN, SB, SLB, SvdW, TM, TA, TM, TL, TPV, TS, TM, TB, UJA, UC, UR, VE, VN, VG, WHR, WAK, WKA, WP, WTC, YAA, YR, YB, YN, YB, ZRdL

## Supplementary Materials

### Methods and Methods

#### Ethics statement

This project relied on sequence data and associated metadata publicly shared by the GISAID data repository and adhere to the terms and conditions laid out by GISAID (*16*). The African samples processed in this study were obtained anonymously from material exceeding the routine diagnosis of SARS-CoV-2 in African public and private health laboratories. Individual institutional review board (IRB) references or material transfer agreements (MTAs) for countries are listed below.

Angola - (MTA - CON8260), Botswana - Genomic surveillance in Botswana was approved by the Health Research and Development Committee (Protocol HPDME 13/18/1), Egypt - Surveillance in Egypt was approved by the Research Ethics Committee of the National Research Centre (Egypt) (protocol number 14 155, dated March 22, 2020), Kenya - samples were collected under the Ministry of Health protocols as part of the national COVID-19 public health response. The whole genome sequencing study protocol was reviewed and approved by the Scientific and Ethics Review Committee (SERU) at Kenya Medical Research Institute (KEMRI), Nairobi, Kenya (SERU protocol #4035), Nigeria – (NHREC/01/01/2007), Mali - study of the sequence of SARS-CoV-2 isolates in Mali - Letter of Ethical Committee (N0-2020 /201/CE/FMPOS/FAPH of 09/17/2020), Mozambique - (MTA - CON7800), Malawi - (MTA - CON8265), South Africa - The use of South African samples for sequencing and genomic surveillance were approved by University of KwaZulu-Natal Biomedical Research Ethics Committee (ref. BREC/00001510/2020); the University of the Witwatersrand Human Research Ethics Committee (HREC) (ref. M180832); Stellenbosch University HREC (ref. N20/04/008_COVID-19); the University of the Free State Research Ethics Committee (ref. UFS-HSD2020/1860/2710) and the University of Cape Town HREC (ref. 383/2020), Tunisia - For sequences derived from sampling in Tunisia, all patients provided their informed consent to use their samples for sequencing of the viral genomes. The ethical agreement was provided to the research project ADAGE (PRFCOVID19GP2) by the Committee of protection of persons (Tunisian Ministry of Health) under the reference (CPP SUD N 0265/2020), Uganda - The use of samples and sequences from Uganda were approved by the Uganda Virus Research Institute - Research and Ethics Committee UVRI-REC Federalwide Assurance [FWA] FWA No. 00001354, study reference - GC/127/20/04/771 and by the Uganda National Council for Science and Technology, reference number - HS936ES) and Zimbabwe (MTA - CON8271).

### Epidemiological and genomic data dynamics

We analyzed trends in daily numbers of cases of SARS-CoV-2 in Africa up to 15 February 2022 from publicly released data provided by the Our World in Data repository for the continent of Africa (https://github.com/owid/covid-19-data/tree/master/public/data) as a whole and for individual countries (*2*). To provide a comparable view of epidemiological dynamics over time in various countries, the variable under primary consideration for **Figure 3** was ‘new cases per million (smoothed)’. To calculate the genomic sampling proportion and frequency for each country for **Figure 1**, the total number of recorded cases at 15 February 2022 was considered, as well as the total length of time for which each country has recorded cases of SARS-CoV-2.

Genomic metadata was downloaded for all African entries on GISAID for the same time period (date of access: 15 February 2022). From this, information extracted from all entries for this study included: date of sampling, country of sampling, viral lineage and clade, originating laboratory, sequencing laboratory, date of submission to the GISAID database. The geographical locations of the originating and sequencing laboratories were manually curated. Sequences originating and sequenced in the same country were defined as locally sequenced, irrespective of specific laboratory or finer location. Sequences originating in one African country and sequenced in another were defined as sequenced within regional sequencing networks. Sequences sequenced in a location not within Africa were labeled as sequenced outside Africa. Sequencing turnaround time was defined as the number of days elapsed from specimen collection to sequence submission to GISAID. Sequencing technology information for all African entries was also downloaded from GISAID on 15 February 2022.

### Primer choice and sequencing outcomes

All SARS-CoV-2 genomes from African countries were retrieved from GISAID (*16*) for submission dates from 1 December 2019 to 28 January 2022 yielding 74 785 entries. Associated metadata for the entries were also retrieved, including collection date, submission date, country, viral strain and sequencing technology. Data on the primers used for the sequencing were requested from investigators and yielded primer data for 13 973 of the 74 785 entries (∼19%). The total N per genome were counted, results from which were then used for genome quality analysis and visualization. Gap locations in the genomes were mapped and visualized compared to the original Wuhan strain (*43*).

### Phylogenetic investigation

All African sequences on the GISAID sequence database (*16*) were downloaded on the 28th of October 2021 (n=48 145). Prior to any phylogenetic inference we performed some quality assessment on the sequences to exclude incomplete or problematic sequences. Briefly, all African sequences were passed through the NextClade analysis pipeline (*44*) in order to identify and exclude: (*i*) sequences missing >10% of the SARS-CoV-2 genome (n=6 495), (*ii*) sequences that deviate by >70 nucleotides from the Wuhan reference strain (n=12), (*iii*) sequences with >10 ambiguous bases (n=165), (*iv*) clustered mutations (n=99), and (*v*) sequences flagged as problematic by NextClade (n=544). Additionally, 492 sequences were removed due to a lack of adequate metadata (i.e. exact sampling date). This produced a final African dataset of 37,787 high quality genotypes for analysis. Due to the sheer size of the dataset we opted to perform independent phylogenetic inferences on the main VOCs (Alpha n=2 365; Beta n=10 809; Delta n=13 911) that have spread on the African continent, as well as a separate inference for all non-VOC SARS-CoV-2 sequences (n=13 252). Due to the discovery of Omicron in November 2021 an additional Omicron dataset was also constructed following the exact quality control measures (n=3 371; 659 did not pass QC; date of access 10th January 2022).

In order to evaluate the spread of the virus on the African continent we aligned the African datasets against a large number of globally representative sequences from around the world. Due to the oversampling of some variants or lineages we performed a random downsampling while retaining the oldest two known variants from each country. In total, 9 335 Alpha, 9 077 Beta, 9 999 Delta, 3 941 Omicron and 16 231 non-VOCs global references were respectively aligned with their African counterparts independently with NextAlign (*44*). Each of the alignments were then used to infer maximum likelihood (ML) tree topologies in FastTree v 2.0 (*45*) using the General Time Reversible (GTR) model of nucleotide substitution and a total of 100 bootstrap replicates (*46*). The resulting ML tree topologies were first inspected in TempEst (*47*) to identify any sequences that deviate more than 0.0001 from the residual mean. Following the removal of potential outliers in R with the ape package (*48*), the resulting ML-trees were then transformed into time calibrated phylogenies in TreeTime (*49*) by applying a rate of 8×10-4 substitution per site per year (*50*) in order to transform the branches into units of calendar time. Time calibrated trees were then visualized along with associated metadata in R using ggtree (*51*) and other packages.

We performed a basic viral dispersal analysis for each of the VOCs (excluding Gamma), as well as for the non-VOC dataset. Briefly, a migration model was fitted to each of the time calibrated tree topologies in TreeTime, mapping the country location of sampled sequences to the external tips of the trees. The *mugration* model of TreeTime also infer the most likely location for internal nodes in the trees. Using a custom python script we could then count the number of state changes by iterating over each phylogeny from the root to the external tips. We count state changes when an internal node transitions from one country to a different country in the resulting child-node or tip(s). The timing of transition events is then recorded which serve as the estimated import or export event. To infer some confidence around these estimates, we performed ten replicates for each of the dataset by random selection from the 100 bootstrap trees. Such phylogeographic methods are always subject to uneven sampling through time (i.e. over the course of the pandemic) and through space (by sampling location). In a previous analysis (*15*) we performed a sensitivity analysis to address some of these issues and found no substantial variations in estimates.

All data analytics were performed using custom python and R scripts and results visualized using the ggplot libraries (*52*).

### Regional and country specific Nextstrain builds

The Nextstrain pipeline (https://github.com/nextstrain/ncov) was used to generate the regional and country-specific builds (*53*). Sequencing data from the rest of the world were included for phylogenetic context based on genomic proximity and even sampling over time. For computational feasibility, ease of interpretation and to balance heterogeneous sampling efforts between countries, genomes were subsampled across time and geography resulting in final builds of ∼6000 genomes for each build.

## Supplementary Figures

**Supplementary Figure S1:**
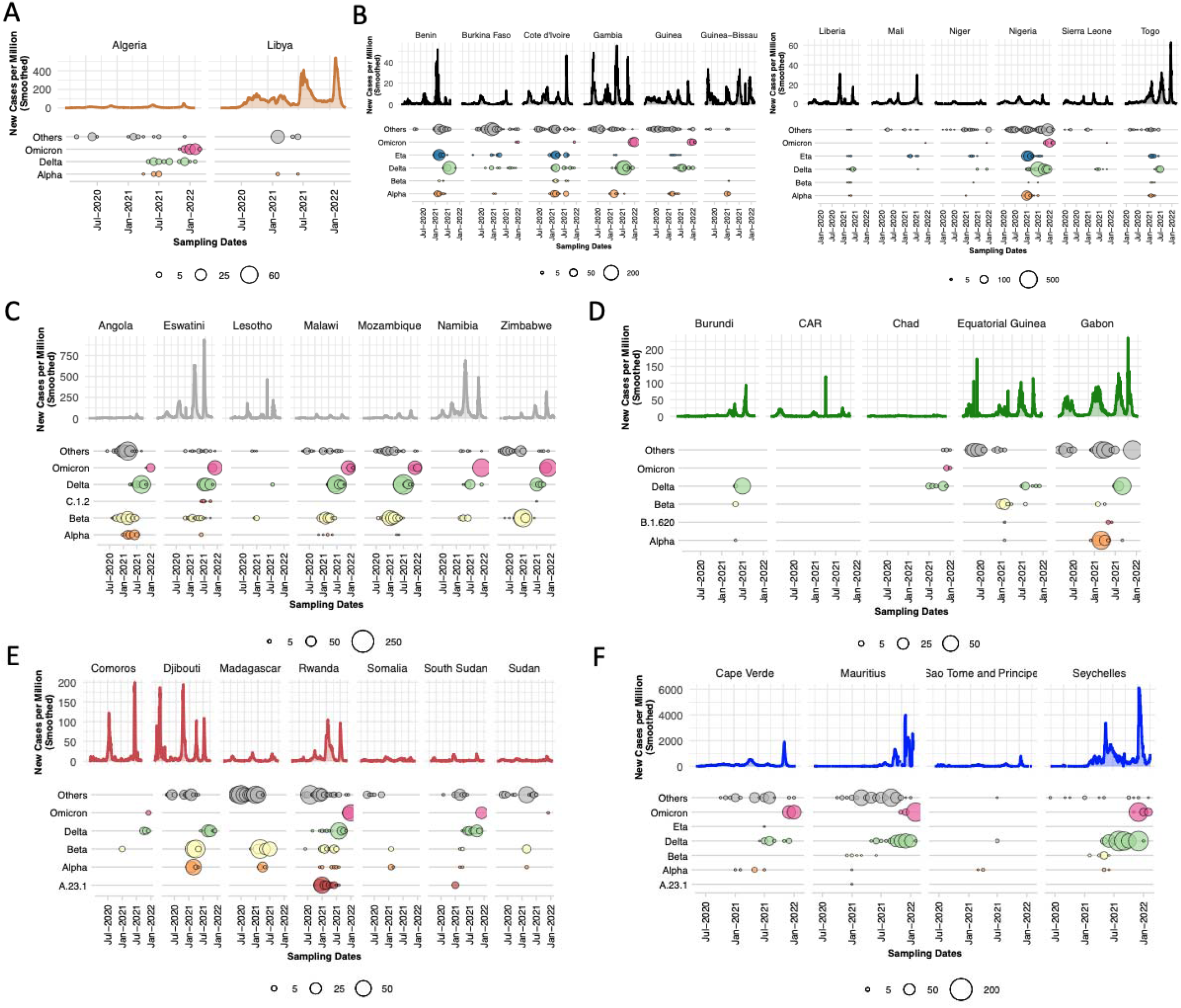
Epidemiological progression of the COVID-19 pandemic in all African countries overlaid with the distribution of VOCs, the Eta VOI and other lineages through time (size of circles proportional to the number of genomes sampled per month for each category). The graphs show a breakdown of new cases per million and monthly sampling of VOCs, regional variant or lineage of interest and other lineages for all African countries not shown in Figure 1, grouped by region: A) North Africa, B) West Africa, C) Southern Africa, D) Central Africa, E) East Africa, F) Cape Verde, Mauritius, Sao Tome and Principe and Seychelles, from the beginning of the pandemic to February 2022.

**Supplementary Figure S2:**
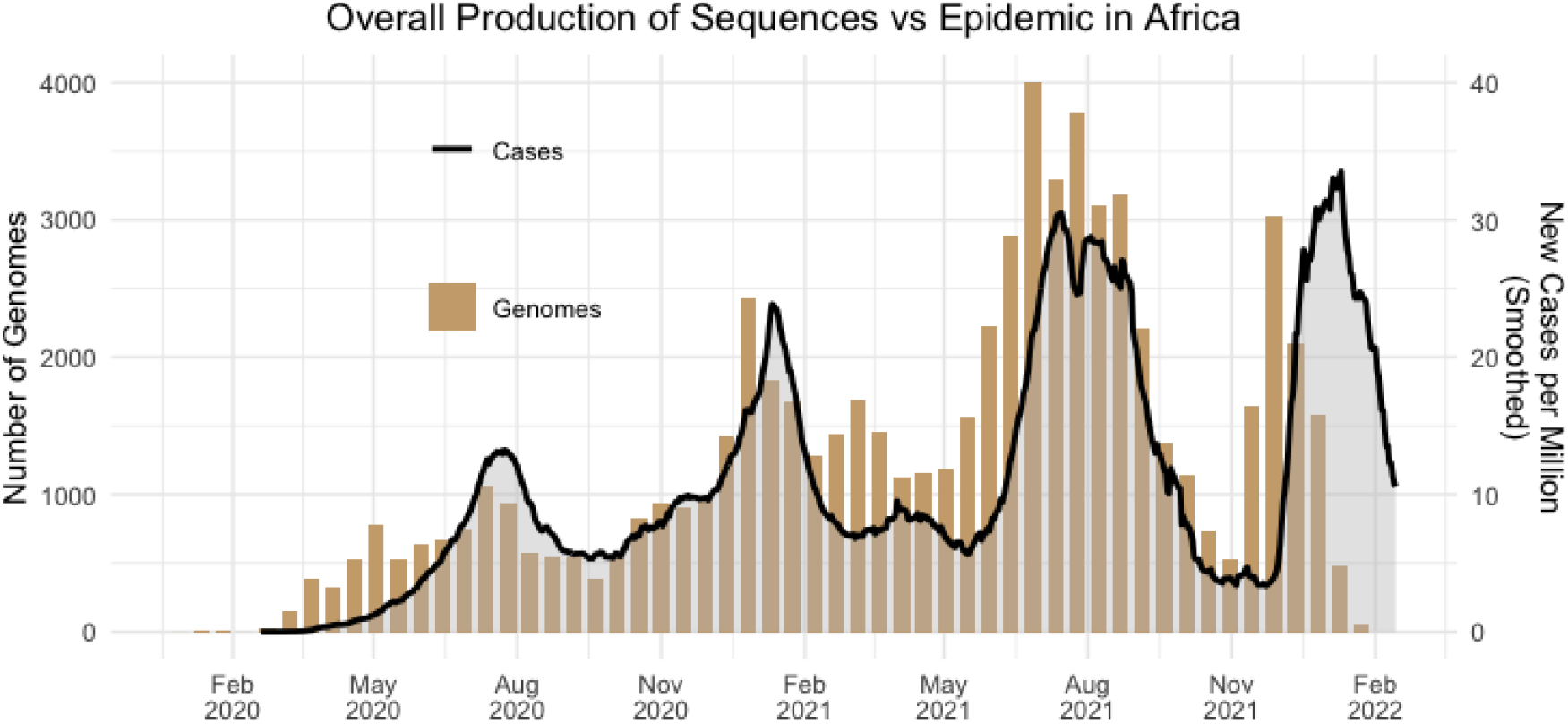
Corresponding progression of genomic sequence production and epidemic size in Africa.

**Supplementary Figure S3:**
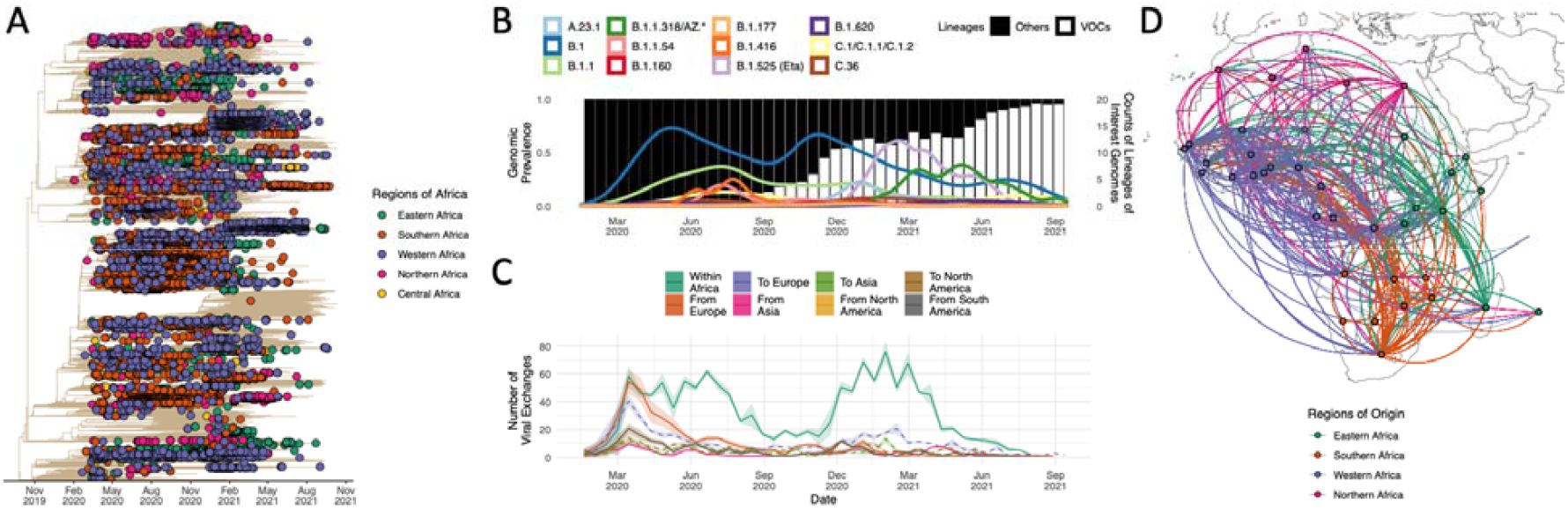
Phylogenetic inference of non-VOC lineages in Africa. A) Maximum-Likelihood timetrees of non-VOC genomes in Africa from the beginning of the pandemic till October 2021 against a global reference with African genomes denoted by tippoint circles coloured by regions of Africa. B) Genomic prevalence of non-VOC (black) vs VOC (white) lineages in Africa overlaid by frequency progressions of some lineages of interest in Africa. C) *Inferred viral dissemination patterns of non-VOC lineages to, from and within the Africa continent from the beginning of the pandemic to October 2021. Introductions and viral transitions within Africa are shown in solid lines and exports from Africa are shown in dotted lines and these are coloured by continent. The shaded areas around the lines represent uncertainty of this analysis from ten replicates. D) Dissemination patterns of the non-VOC lineages within Africa, from inferred ancestral state reconstructions, annotated and coloured by region in Africa. The countries of origin of viral exchange routes are also shown with dots and the curves go from country of origin to destination country in an anti-clockwise direction*.

**Supplementary Figure S4:**
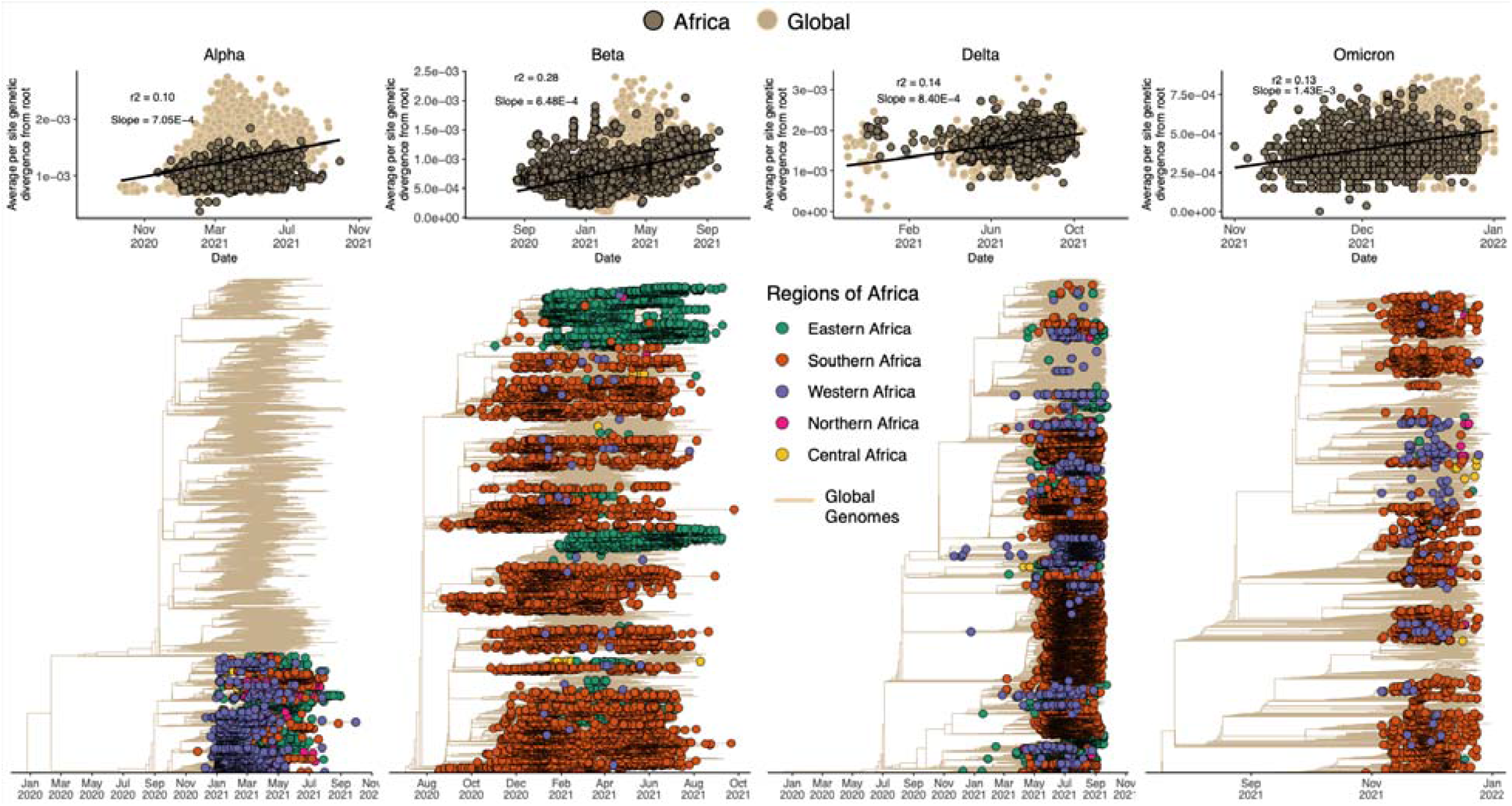
Phylogenetic inference of VOCs in Africa. Top - Molecular clock evolution. Bottom - VOC-specific Maximum-Likelihood timetrees with African genomes denoted by tippoints coloured by regions of Africa.

**Supplementary Figure S5:**
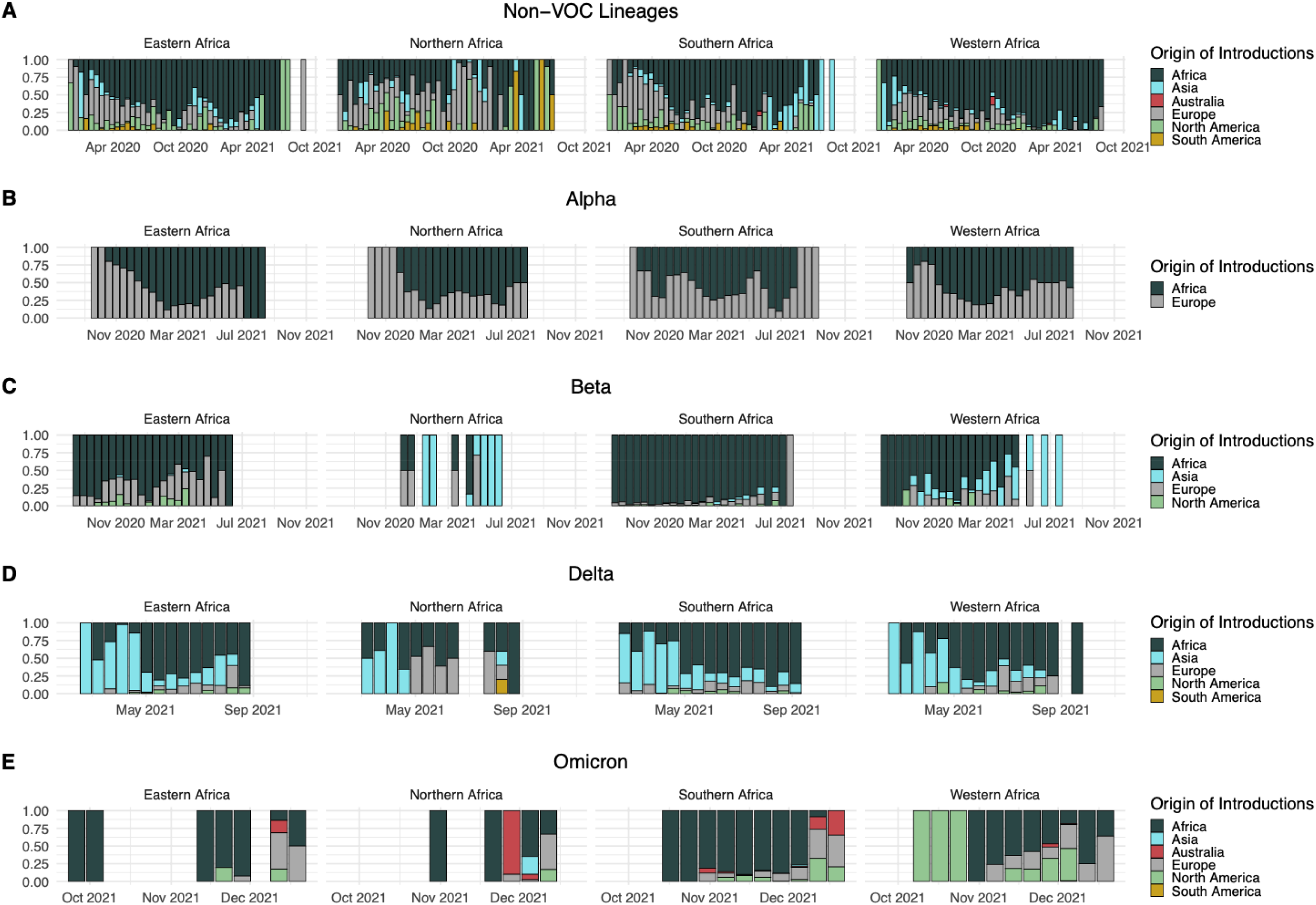
Patterns of viral importations into different regions of Africa showing the proportions of introductions into Eastern, Northern, Southern and Western Africa attributed to specified origins for A) non-VOC lineages, B) Alpha VOC, C) Beta VOC, D) Delta VOC, and E) Omicron VOC during the relevant time periods.

**Supplementary Figure S6:**
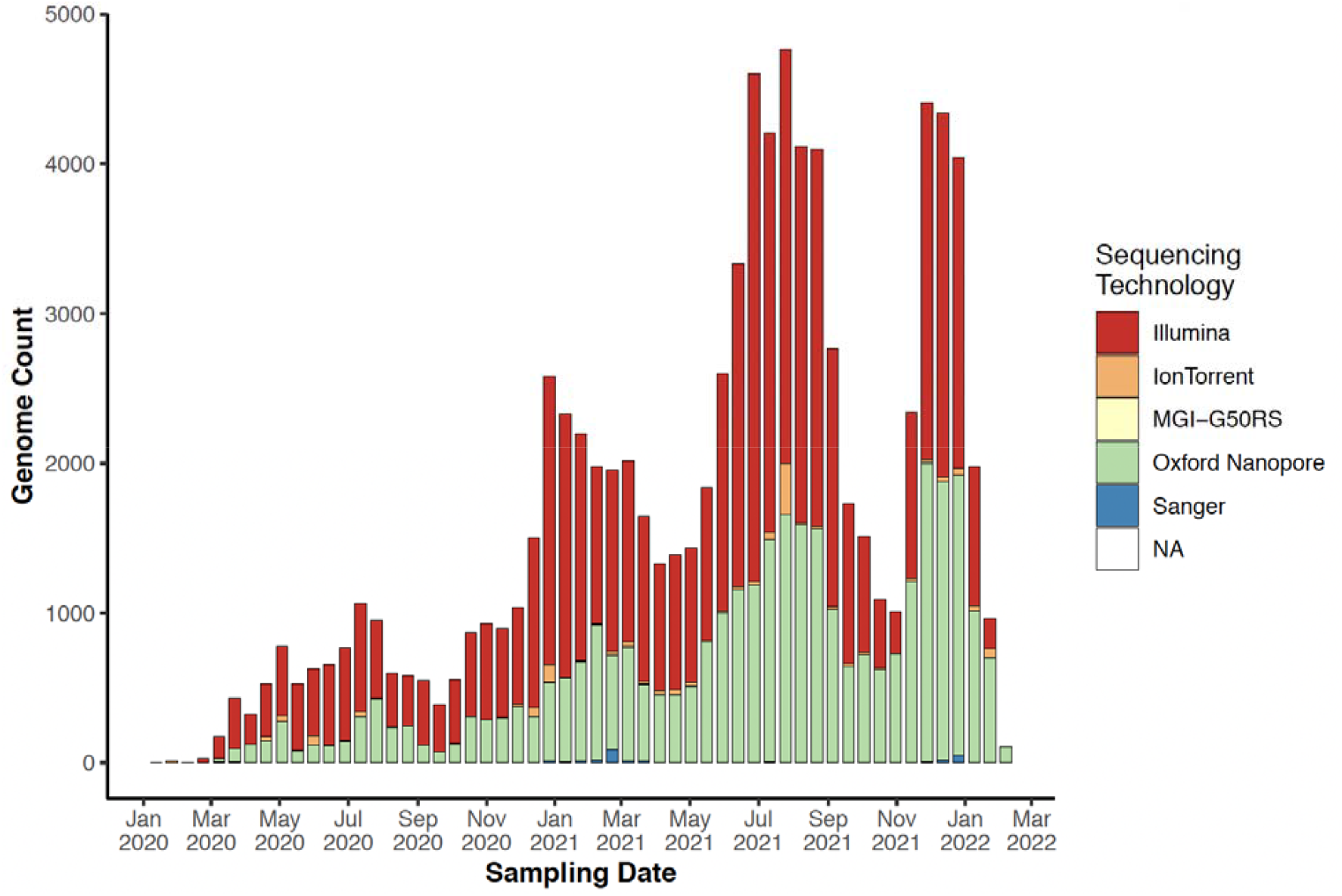
The progression of sequencing technologies used for SARS-CoV-2 sequencing in Africa

**Supplementary Figure S7:**
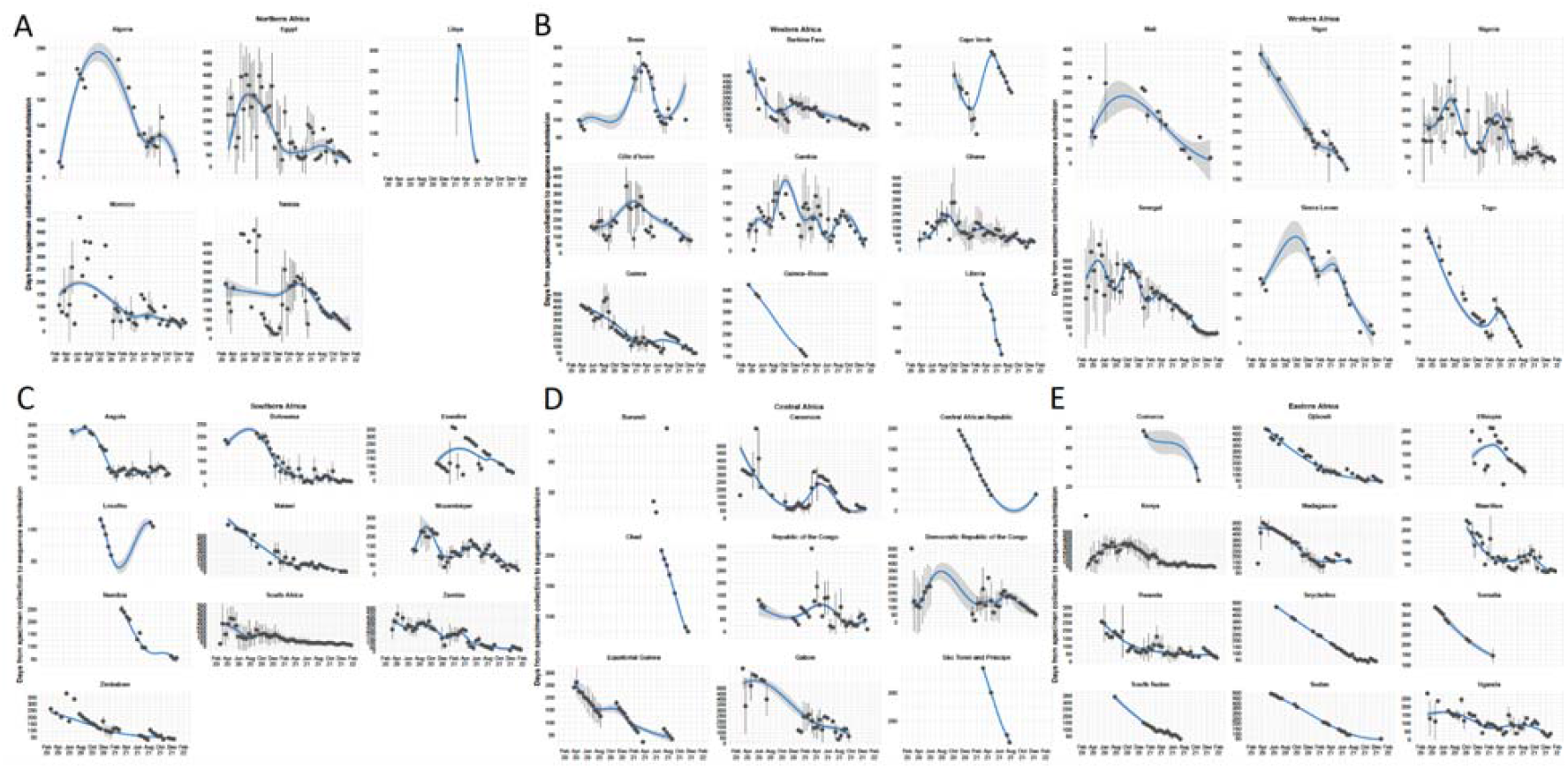
Sequencing turnaround time progression per country in each region of Africa in A) North Africa, B) West Africa, C) Southern Africa, D) Central Africa, and E) East Africa

**Supplementary Figure S8:**
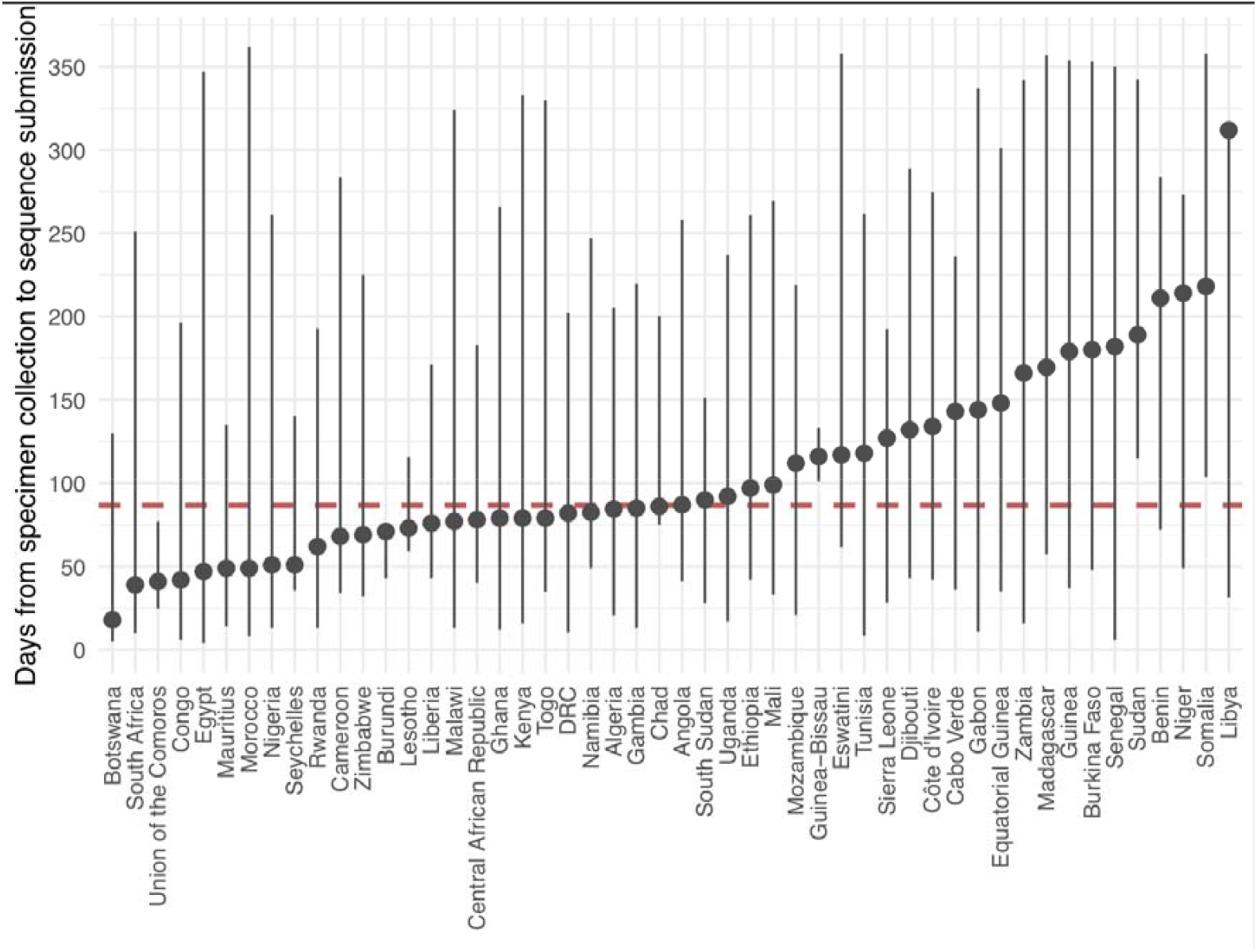
Sequencing turnaround time average by country. The circles indicate the mean number of days between specimen collection and sequence submission to GISAID and the bars indicate the distribution days.

**Supplementary Figure S9.**
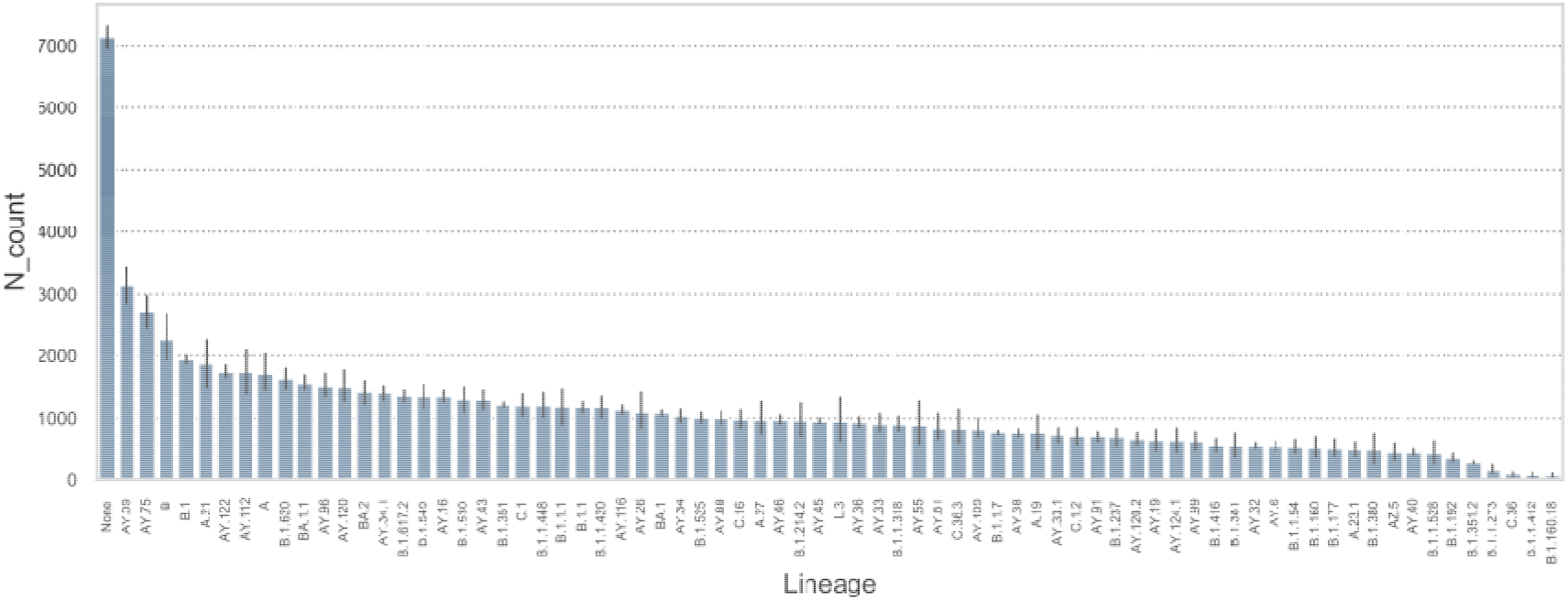
Gapped genomes by lineage. The mean N data were stratified by SARS-CoV-2 lineages to investigate if there were lineage-specific patterns in the genome gaps, an indirect measure of primer mismatch. For lineages present in at least 100 times in the Africa 75K genome data, there was a modest difference in N content across the lineages. Lineages that returned no classification with Pangolin (“None”) showed the highest N content, which is not surprising as the high N content was probably the basis for the failed classification. The more recent lineages Delta (e.g. AY.39, AY.75) and Omicron (BA.1.1, BA.2) also showed higher N content. There were however a number sequences classified into more basal lineages (e.g. B, B.1) and it is possible that the basal lineage accumulated genome sequences that, because of their N content, failed to return a more specific classification.

**Supplementary Table S1:** NextStrain builds of the five major geographical regions in Africa. Each build focuses on a specific region within Africa and includes sequences from outside the region and the continent to place the regional sequences into context of the global pandemic. All the data used in the builds are publicly available on GISAID and are maintained and updated by the Africa CDC in collaboration with the NextStrain team on a weekly basis.

| Region | Regional Builds |
| --- | --- |
| Central African Region | https://nextstrain.org/groups/africa-cdc/ncov/central-africa |
| Eastern African Region | https://nextstrain.org/groups/africa-cdc/ncov/eastern-africa |
| Northern African Region | https://nextstrain.org/groups/africa-cdc/ncov/northern-africa |
| Southern African Region | https://nextstrain.org/groups/africa-cdc/ncov/southern-africa |
| Western African Region | https://nextstrain.org/groups/africa-cdc/ncov/western-africa |

**Supplementary Table S2:**
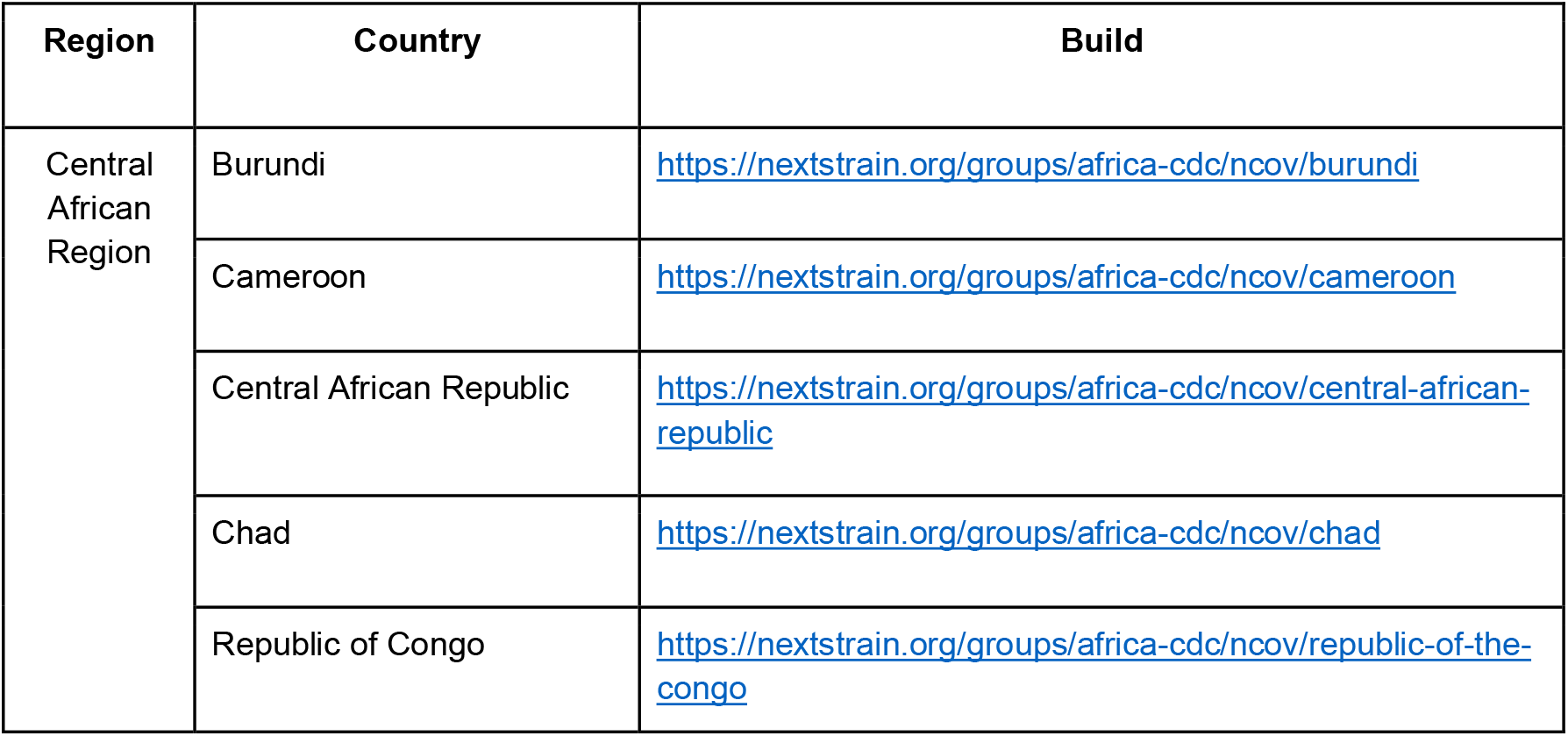
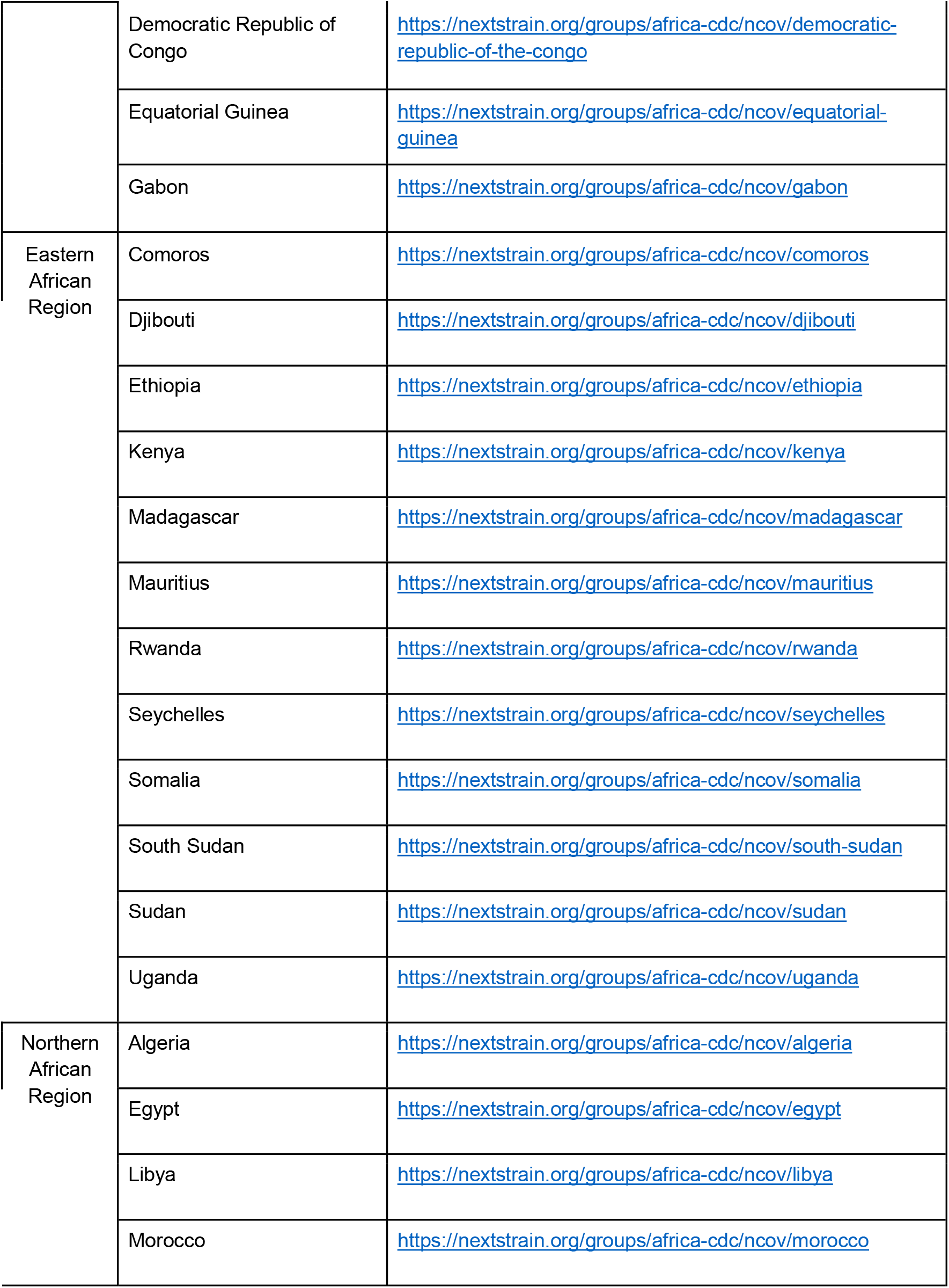
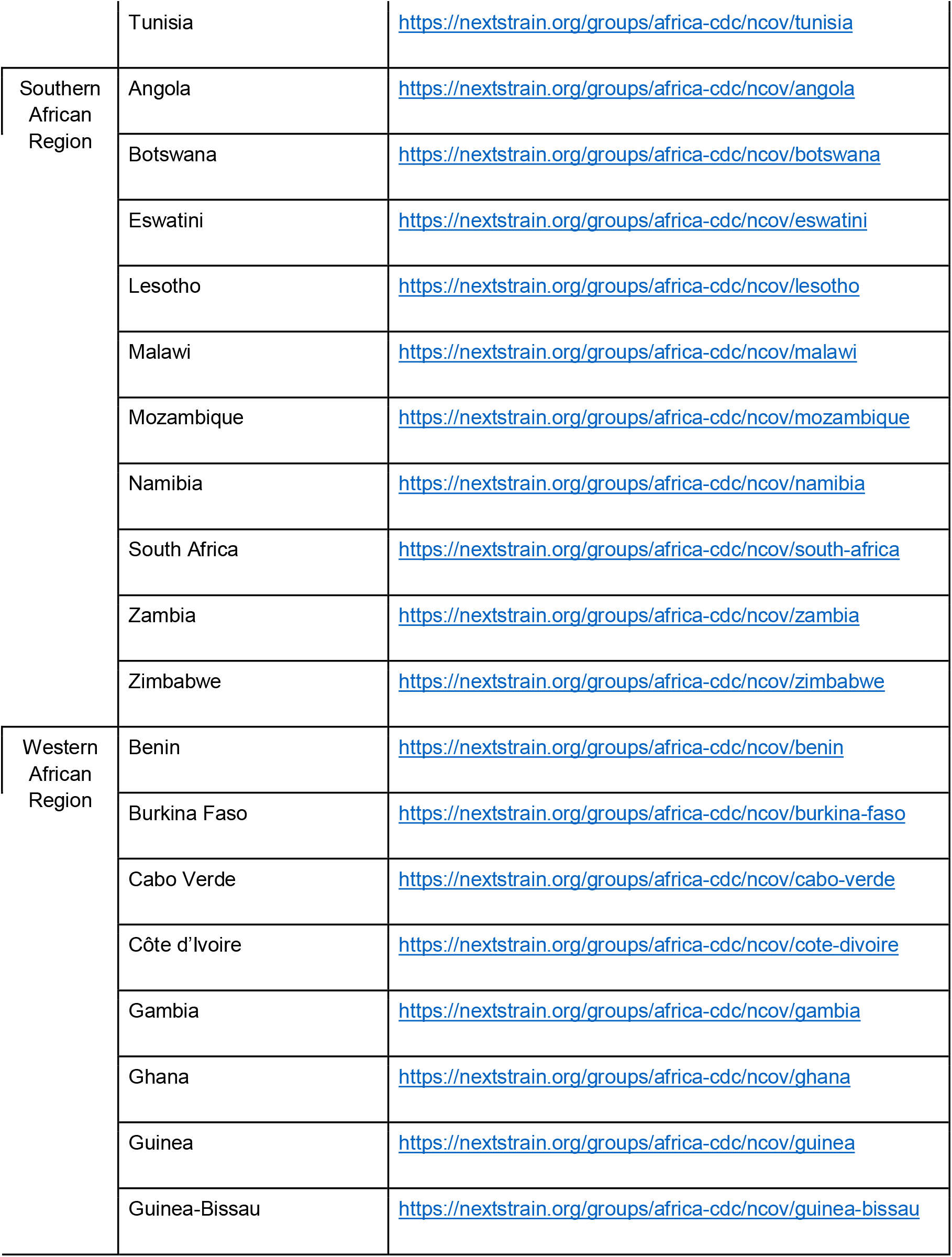
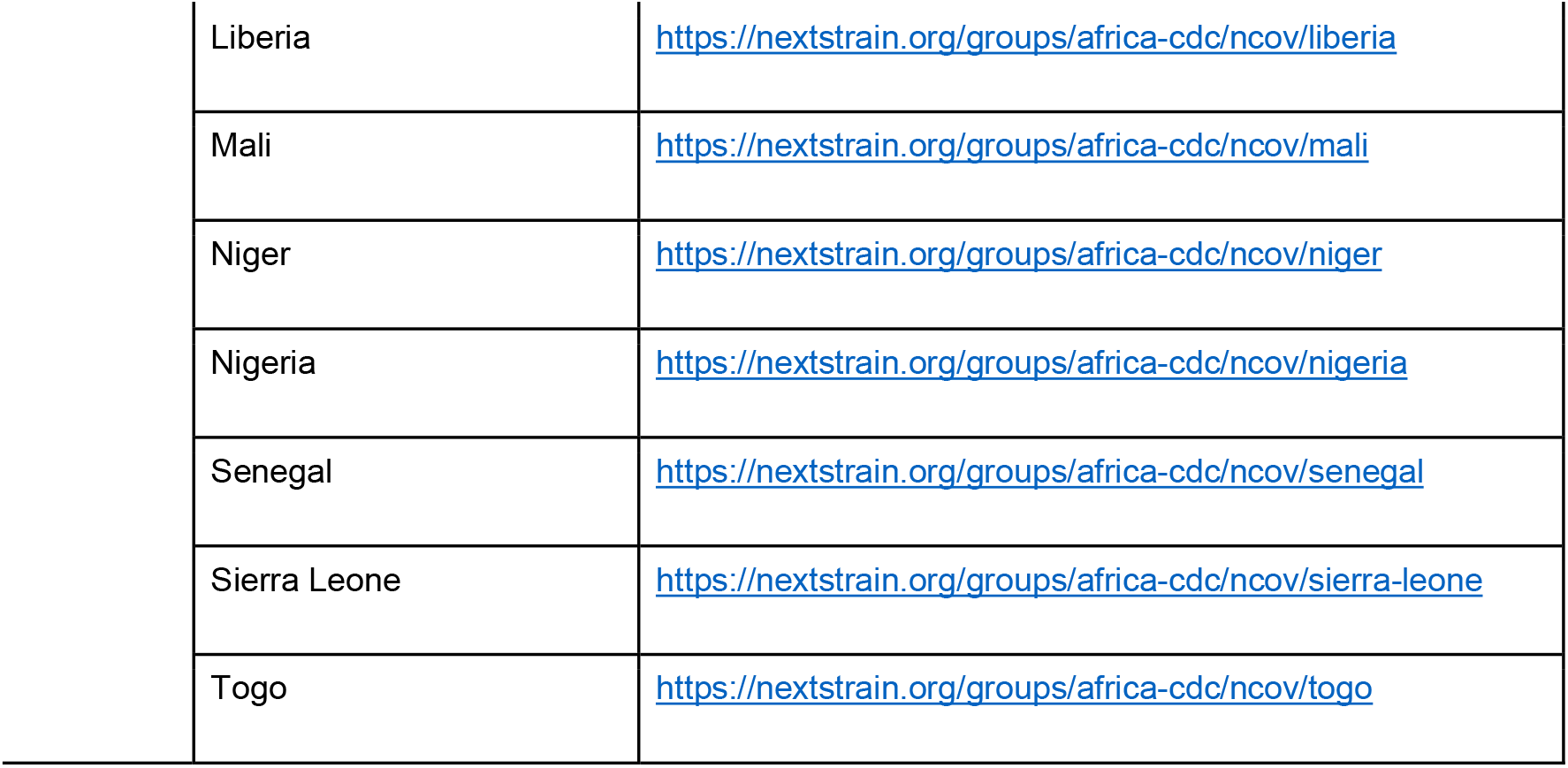
Individual country specific NextStrain builds from the African continent. Each build focuses on a specific country within Africa and includes sequences from the rest of the world in order to place the country’s sequences into context of the global pandemic. All the data used in the builds are publicly available on GISAID and are maintained and updated by the Africa CDC in collaboration with the NextStrain team on a weekly basis.

## Supplementary Author List

Aaron L. Shibemba, Abasi Ene Obong, Abayomi FADEYI, Abbad Anas, Abd Elazeez Shabaan, Abd Monaem Adel, Abd Moniem Ain Shoka, Abdelhamid W., Abdelilah Laraqui, Abdelkader Laatiris, Abdelkrim Meziane Bellefquih, Abdellah Faouzi, Abdelmoulah F., Abdelomunim Essabbar, Abderrahmane Bimouhen, Abderraouf Hilali, Abdo I., Abdou Padane, Abdoul Karim Sangaré, Abdoul Karim Soumah, Abdoulaye Djimde, Abdoulaye Toure, Abdoulie Kanteh, Abdulla Bashein, Abdullah Salama, Abe G. Abias, Abebe Genetu Bayih, Abel Abera Negash, Abel Lissom, Abid N., Abla A. Konou, Abo Shama, Abosede O., Abouelnaga S., Abraham Ali, Abraham Kwabena Anang, Abraham Tesfaye, Adam K. Khan, Adamu Tayachew, Adane Mihret, Adba Alfatih AlEmam, Adede Hawi, Adesegun A., Adey Feleke Desta, Adib Ghassan, Adjaratou Traoré, Adjiratou Aissatou B. A., Adodo Sadji, Adrian Egli, Adriano Mendes, Adugna Abera, Adul Candé, Afaf Alaoui, Afonso Pedro, Agbodzi B., Ageez A.M., Ahidjo Ayouba, Ahmed E Kayed, Ahmed El-Taweel, Ahmed Elsayed, Ahmed F. Gad, Ahmed Fakhfakh, Ahmed Kandeil, Ahmed M., Ahmed Mostafa, Ahmed O. S., Ahmed Reggad, Ahmed Taha, Ahyong Vida, Aicha Bensalem, Aida Sivro, Aissam Hachid, Ajili F., Ajogbasile F.V., Akim A. Adegnika, Akoele Siliadin, Akwii Patience Natasha, Aladje Balde, Alan Lemtudo, Alaoui Sanaa-amine, Alaruusi A. M., Alassane Ouro-Medeli, Albert Nyunja, Alberto Rizzo, Alemseged Abdissa, Alemu Tike Debela, Alessandro Mancon, Alessandro Marcello, Alexander Goredema, Alexander Greninger, Alexis Ndjolo, Alexis Niyomwungere, Alfredo Mari, Alfredo Mayor, Ali M.A., Ali Zumla, Alia Ben Kahla, Alia Grad, Alice Kabanda, Alie Tia, Alimou Camara, Alimuddin Zumla, Alle Baba Dieng, Almoustapha I. Maiga, Amadou Alpha Sall, Amadou Daou, Amal Naguib, Amal Souiri, Amal Zouaki, Amalou Ghita, Amandine Mveang-Nzoghe, Amariane Koné, Amariane M. M. Koné, Ambroise Ahouidi, Amel Benyahia, Amel Nagiub, Amer K. E., Ameyo Dorkenoo, Amina Barkat, Aminata Dia, Aminata Mbaye, Aminata Mboup, Aminata Sileymane Thiam, Amine Idriss Lahlou, Amira Suliaman Wadi, Amivi Ehlan, Amujal Marion, Amuri Aziza, Amy Strydom, Anass Abbad, Anatole Nkeshimana, Andargachew Mulu, Anderrahmane Maaroufi, Andrea E. Luquette, Andreas Shiningavamwe, Andres Moreira-Soto, Andrew Azman, Andrew J. Bennett, Andrew Tarupiwa, Anga Latifa, Ange Badjo, Angel Angelov, Angela Brisebarre, Angela M. Detweiler, Angoune Ndong, Anja Werno, Anna Julienne Selbe NDiaye, Anna-Lena Sander, Annajr B., Annamaria D’Aprile, Anne van der Linden, Annemiek van der Eijk, Annette Erhart, Anou M. Somboro, Anoumou Dagnran, Anthony Ahumibe, Anthony Levasseur, Antje van der Linden, Antoine Dara, Anu Jegede-Williams, Aouni M., Arjarquah A., Arlene Uwituze, Arlo Upton, Armel Poda, Arsène Somé, Arsène Zongo, Arsenia Massinga, Asare K.M., Ashaba Fred Katabazi, Asma Ferjani, Assane Dieng, Astou Gaye-Gaye, Atiga N., Atsbeha G. Weldemariam, Augustina Arjaquah, Auld A., Awa Ba-Diallo, Awatef ElMoussi, Awunyo Sena, Aya Mohamed, Ayman Farghaly, Ayo-Ale B., Ayoade F., Ayola Akim Adegnika, Ayong More, Ayorinde Babatunde James, Azami Nawfel, Azaria Diergaardt, Azuka Patrick Okwuraiwe, Babafemi O. Taiwo, Baboo S. B., Bahadoor B. S., Bahnassy A.A., Bakary Sanyang, Bakry U., Bamba Fatoumata Touré, Bamidele Iwalokun, Banda R, Bane S., Bankole Johnson, Barada Cisse, Barbra Murwira, Bas Oude Munnink, Batchi-Bouyou Armel L., Batra R., Belayachi Lamiae, Belkhir A.B., Ben Ayed I., Ben Morton, Ben Moussa, Ben Wulf, Bénédicte Ndeboko, Benhida Rachid, Benjamiin B. Lindsey, Benjamin H. Foulkes, Benjamin Hounkpatin, Benjamin Selekon, Bensaid M., Bernard Ssentalo Bagaya, Bert Vanmechelen, Bertrand Lell, Beth Mutai, Bethlehem Adnew, Beuty Makamure, Bighignoli B., Birahim Piere Ndiaye, Bishwo N. Adhikari, Bitrou S.M., Blaise MBoringong Akenji, Bode Shobayo, Boitumelo Zuze, Bonifacio Manguire Nlavo, Bouchra Belfquih, Bouchra Boujemla, Bouna Yatassaye, Bouzidi Aymane, Brian Andika, Bright K. Yemi, Bronwyn Kleinhans, Bruna Galvao, Bubacar Delgado Pinto Embaló, Bulelani Manene, Camille Capel, Campbell Anu, Carine Tchibozo, Carla Madeira, Carlos Cortes, Carniel Elisabeth, Carol Kifude, Carolle Yanique Tayimetha, Catherine E. Arnold, Catherine Okoi, Cecilia Waruhiu, Celestin Godwe, Celestina Obiekea, Celine Nkenfou, Chabuka L. B., Chakib Nejjari, Chambaro H., Chanda D., Changula K., Charifa Drissi Touzani, Charles Kayuki, Charles Nyagupe, Charoute Hicham, Chaselynn M. Watters, Chawech H., Cheikh Sokhna, Chenaoui Mohamed, Chiamaka Nwuba, Chilufya C., Chimaobi Chukwu, Chinyere Anyika, Chipimo P.J., Chitanga S., Chitenje M., Chiwaula M. J., Chouati Taha, Chris Mansell, Christelle Butel, Christian A. Devaux., Christian Drosten, Christian Ranaivoson, Christian Utpatel, Christophe Malabat, Chtourou A., Chukwu G., Claudia Daubenberger, Clement G. Kakai, Clement Masakwe, Clotaire Donatien Rafai, Collins Chenwi, Collins M. Misita, Collins Muli, Corine H. GeurtsvanKessel, Corinne Maufrais, Coulibaly Mbegnan, Crawford M., Cristina M. Tato, Cyrus Yiaba, D’Amore N., Dabiri Damilari, Dalia Ramadan, Damena D., Daniel Ouso, Daniela Pisanelli, Danilo Licastro, David Hammer, David Nieuwenhuijse, David Patrick Kateete, David Wilkinson, Davy Leger Mouangala, Dawit Hailu Alemayehu, Dawood R.M., De Sanctis R., Deborah N. A. Mettle, Demba Koita, Dennis Kenyi Lodiongo, Dennis Laryea, Department of Virology, College of Medicine, University of Ibadan, Dereje Leta, Dersi Noureddine, Desire Takou, Dessalegn Abeje Tefera, Dhwani Batra, Dia Ndongo, Diab A., Diabou Diagne, Diane A. Mabika, Diané Bamourou, Dianke Samaté, Diarra B., Didier Raoult, Dieudonne Mutangana, Dineo Emang Tshiamo. Gape Nyepetsi, Diosdado Odjama Nseng Ada, Djimabi Salah, Donatella Cedola, Doris Harding, Dorothy Yeboah-Manu, Dossou Ange, Dowbiss Meta Djomsi, Dragana Drinkovic, Draper C., Dumbuya Foday Sahr, Ebenezer Odewale, Eclou Sedjro, Edang-Minko Armand, Edgar Kigozi, Edith Koskei, Edith Nkwembe, Edmilson F. de Oliveira Filho, Edmira Maria da Costa, Edward Kiritu, Efrem S. Lim, Egon A. Ozer, Egyir B., Ehab Abdelkader Abuelenein, Eitel Mpoudi Ngole, El Aliani Aissam, El Ansari Fatima Zahra, El Hamouchi Adil, El Oualid Abdelmjid, El-Shaqnqery H., El-Zayat M., Elamin Abualas, Elannaz Hicham, Elargoubi A., Eleni Kidane, Elham Rizgalla, Elhoseny M.M., Elhosieny F.W., Elisabeth Carniel, Elizabeth Nyakarungu, Elkhateeb S.M., Elmostafa Benaissa, Elmostafa El Fahime, Elouanass M., Elsissy M.H., Elvyre Mbongo-Nkama, Elwaleed M. Elamin Sara A.I Latif, Emame Edu, Emanuele Orsini, Emilio Skarwan, Emily K. Stefanov, Emmanuel Nasinghe, Emmanuel Nepolo, Emmanuel Ogunbayo, Emmanuelle Munger, Emmanuelle Permal, Emna Gaies, Ennibi H., Ennibi Khalid, Erasmus Kotey, Erasmus Smit, Eric A. Lelo, Eric Adu, Eric Delaporte, Eric Katagirya, Eric Muthanje, Erica Luis Maria Magalhães, Ernest Asiedu, Esemu Livo, Esperance Umumararungu, Esther Chitechi, Esther Omuseni, Etoundi Mballa Alain, Evelyn Bonney Quansah, Evelyn Y. Bonney, Ezzelarab M.H., Faatu Cassama, Fabien Roch Niama, Fabio Arena, Faggioni G., Fahsbender E., Faith Sigei, Faouzi Abdellah, Farah Jouali, Farawyla H., Farida Hilali, Fatima Chgouri, Fatima El Falaki, Fatima Zahra El ansari, Fatma Ebied, Fatna Bssaibis, Fattah Al Onifade, Faye Ousmane, Fayez Ahmed Khardine, Fayez Khardine, Fedorov A.V., Fekadu Alemu, Fekkak Jamal, Fengming Sun, Fetouma Doudou, Fikry A.E., Fillo S., Fiorenza Bracchitta, Firmin Kaboré, Fki-berrajah, Flora Donati, Florence Fenollar, Foday Sahr Samuel Sorie, Folarino O., Folorunsho, Francisca Muyembe-Mawete, Francisco Malagon, François Kiemdé, Franklin Asiedu-Bekoe, Franklyn Egbe Nkongho, Frédéric Lemoine, Frederick Tei-Maya, Freitas R. H., Fuh-Neba T., Gaaloul I., Gabriel Kabamba, Gadissa Gutema, Gaies Emna, Galadima Gadzama, Galal Mahmoud, Garba Ouangole, Gargouri S., Garry R., Gary McAuliffe, Gathii Kimita, Gédéon Prince Manouana, Gelanew Tesfaye, George Awinda, George B. Kyei, George Michuki, Georgelin Nguema Ondo, Gerhard van Rooyen, Gert Marais, Getachew Abichu, Getachew Tesfaye Beyene, Getachew Tollera, Getnet Hailu, Ghamaz Hamza, Ghazi Kayali, Ghizlane El Amin, Gibson Mhlanga, Gilbert Kibet, Gildas Hounkanrin, Giordani F., Gizaw Teka, Goldstone R., Gomaa C., Gomaa Mokhtar, Gora Lo, Gosnell B. I., Gottberg A., Grace Angong Belournou, Grace Oni, Grace Vincent, Guedi Ali Barreh, Guillermo Garcia, Guohong Deng, Guseva N. P., Guy Paterne Malonga Mbembo, Guy Stéphane Padzys, Gwayi S., Habiba Ben Romdhane, Habiba Naija, Haby Diallo, Hadad A., Hafez M.M., Hafsia Ladhari, Hagar Elshora, Hailu Dadi, Hajjaji Mohammed, Hakima Kabbaj, Hala Hafez, Halafawy A., Halidou Tinto, Halimatou Diop Ndiaye, Hamadi Assane, Hamdani T.N., Hamdy M.S., Hamidah Namagembe, Hammad M., Hammami A., Hamza Gharmaz, Hana Sofia Andersson, Hanae Dakka, Hanen El Jebari, Hany K. Soliman, Harmak Houda, Harvey R., Hasnae Benkirane, Hassan Aguenaou, Hassan Ihazmad, Hassan R., Hassan W., Hatem Ahmed, Heitzer Sogodogo, Helena Seth-Smith, Helisoa Razafimanjato, Hellen Koka, Hemlali Mouhssine, Henry Mwebesa, Hermes Perez, Herve Christian Paho Tchoudjin, Hicham Elannaz, Hicham Oumzil, Hilda Opoku Frempong, Hinson Fidelia, Hoda Ezz Elarab, Hong Xie, Houda Benrahma, Housna Arrouchi, Houssem Guedouar, Hu Luo, Hubert Bassène, Huiqing Si, Ian Goodfellow, Ibrahima Guindo, Ibrahima Halilou, Idil Salah Abdillahi, Idriss-Amine Lahlou, Idrissa Diawara, Ifunanya Egoh, Ige F.A., Iknane A.A., Ikram Bnouyahia, Ikram Omar Osman, Ilhem Boutiba-Ben Boubaker, Iman Abdillahi Hassan, Iman Foda, Imane Smyej, Imen Kacem, Imen Mdini, Imen Mkada, Inacio Mandomando, Iñaki Comas, Ines Mdini, Inglês L., Innocent Mudau, Ireoluwa Yinka JOEL, Irina Chestakova, Irving Cancino, Isaac Phiri, Ismael Pierrick Mikelet Boussoukou, Ismail A., Israel Osei-Wusu, Issaka Maman, Ivan Barilar, Ivy A. Asante, Izuwayo Gerard, Jaafar Heikel, Jacob Souopgui, Jacques Marx, Jalal D., Jalal Nourlil, Jalil El Atar, Jalila Ben Khelil, Jalila Rahoui, Jamal Fekkak, James Mutisya, James Ussher, Jan Felix exler, Jane MaCauley, Janet Majanja, Jannoo N., Jarra Manneh, Jasmin Scharnberg, Jasmin Schlotterbeck, Jasper Chimedza, Jawad Bouzid, Jean Claude Djontu, Jean Maritz, Jean-Claude Makangara Cigolo, Jean-louis Monemou, Jeanne d’Arc Umuringa, Jeremy Delerce, Jerome Nkurunziza, Jesse Addo Asamoah, Jill Sherwood, Jing Wang, Joan Marti-Carerras, Joe K. Mutungi, Joe Mutungi, Joel Fleury Djoba Siawaya, Joel Koivogui, Joep de Ligt, John Njuguna, John Rumunu, John Tembo, John Waitumbi, Johnson A. Adeniji, Jonathan Rigby, Jorn Hellemans, Joseph Fokam, Joseph Francis Wamala, Joseph L. DeRisi, Joseph Makhema, Joseph Ojonugwa Shaibu, Joseph Oliver-Commey, Josh Freeman, Josiah Ayoola Isong, Josphat Nyataya, Joy Ayoola, Joyce Appiah-Kubi, Judd F. Hultquist, Jude Gedeon, Judith Sokei, Julia Howard, Julia Schneider, Julian Campbell, Juliet Elvy, Juma John H. M., Jumaa A.B., Justin Lee, Justin Lessler, Justin O’Grady, Kaba Kourouma, Kais Ghedira, Kalantar K., Kamela Mahlakwane, Kamoun S., Kangwa Mulonga, Kapata P.C., Kapaya F., Kapin’a M., Karray Hakim H., Kasambara W., Kasmi Yassine, Kassahun Tesfaye, Kathleen Subramoney, Katyshev A.D., Kayeyi N., Kayla Barnes, Kayla Delaney, Kazorina E.V, Kedumetse Seru, Keita S., Keith Durkin, Keith R Jerome, Keke K. René, Kena Swanson, Kenneth K. Maeka, Keren Okyerebea Attiku, Kevin Sanders, Kevine Zang Ella, Keyru Tuki, Khabab A. Elhag, Khadim Gueye, Khaled Amer, Khalid ENNIBI Mostafa, Kharat N., Khumalo Z., Kilian Stoecker, Kim L., Kimberly A. Bishop-Lilly, Kimotho J., Kitane Driss Lahlou, Kofi Bonney, Kokou Tegueni, Kolawole Wasiu WAHAB, Kolomoets E.V., Komal Jain, Kominist Asmamaw, Komlan Kossi, Kondwani Jambo, Kouriba Dürr, Kra Ouffoué, Krasnov Y.M., Krishna Kumar Kandaswamy, Kristian Andersen, Kritsky A.A., Kumbelembe David, Kutlo Macheke, Kutyrev V.V., Kwenda S., Kwitaka Maluzi, Kwok Lee, Kyle A. Long, Kyra Grantz, Lacy M. Simons, Laetitia Serrano, Lagos State Government, Laila Elsawy, Laila Sbabou, Lallepak Lamboni, LaRinda A. Holland, Lasata Shrestha, Lassana Sangaré, Latifa Anga, Lauren Jelly, Laurien Hoornaert, Le Thi Kieu Linh, Legodile Kooepile, Leigh-Anne MC Intyre, Léon Mutesa, Leona Okoli, Léopold Ouedraogo, Lesego kuate-lere, Leta D., Letaief A., Liboro G., Lilian Kanjau, Lin L., Linda Boatemaa, Linda Houhamdi, Lipkin W. Ian, Lista F., Liwewe M.M., Lloyd Mulenga, Logan J. Voegtly, Loide Shipingana, Loris Micelli, Lorreta Kwasah, Loubna Allam, Louise Lefrançois, Loukman Salma, Lucas N. Amenga-Etego, Ludivine Brechar, Ludovic Mewono, Luis A. Estrella, Lusia Mhuulu, Lwanga Newton, M. Ulrich, M.T. Mogotsi, Maaroufi Abderrahmane, Mad W., Madi W.Y., Madlen Stange, Magdeldin S., Maher Kharrat, Mahlangu B, Mahmoud Shehata, Mahrous N., Maida A, Makhtar Camara, Makori T., Malama K., Malena S. Bestehorn-Willmann, Malolo I., Mamadou Beye Cheikh Ibrahima Lo, Mamadou Bhoye Keita, Mamadou Saliou Bah, Mamadou Saliou Sow, Mamoudou Harouna Djingarey, Mamoudou Maiga, Manal Hamdy Elsaid, Manal Hamdy Zahran, Mandiou Diakite, Manel Ben Sassi, Mangombi Pambou, Manickchund N., Manoli Torres Puente, Manraj S. S., Mansour T., Maowia M. Mukhtar, Marcel Tongo, Marchoudi Nabila, Margaret Mills, Maria Artesi, Maria Pia Patrizio, Maria Rita Gismondo, Maria Rosaria Lipsi, Mariam Kehinde SULAIMAN, Mariama Kujabi, Marie Amougou, Marie Claire Okomo, Marie Madeleine Chabert-Consen, Marie-Astrid Vernet, Marie-Pierre Hayette, Mariem Gdoura, Marijke Reynders, Marion Barbet, Marion Koopmans, Marjan Boter, Markos Abebe, Markus H. Antwerpen, Marouane Melloul, Martin Maidadi Foudi, Martine Peeters, Marvin Hsiao, Mary DeAlmeida, Mary Lalemi, Mary-Ann Davies, Masahiro K., Masse Sambou, Mathabo M., Mathew D. Parker, Mathias C. Walter, Mathur H., Matt Blakiston, Matt Storey, Matthew Bates, Matthew Rogers, Matthias Pauthner, Maud Vanpeene, Maurizio Margaglione, Max Bloomfield, May Abdelfattah, May Sherif Soliman, Mbengué Fall, Mdlalose K., Meei-Li Huang, Mehta S., Mélanie Albert, Melchior A. Joël Aïssi, Méline Bizard, Merabet Mouad, Meriem Laamarti, Messanh Douffan, Mhalla S., Michael Addidle, Michael Marks, Michael Nagel, Michael V. Deschenes, Micheala Davids, Michelle Balm, Michelle Lin, Michelle Tan, Mihrete A., Mikhail Olayinka BUHARI, Milanca Agostinho Cá, Mildred Adusei-Poku, Milkah Mwangi, Mina Kamel, Miranda J., Mireille Prince-David, Miriam Eshun, Misaki Wayengera, Mitali Mishra, Mjid Eloualid, Mly Abdelaziz Elalaoui, Mnguni A., Mnyameni F., Mogomotsi Matshaba, Mohale T., Mohamed Abdel-Salam Elgohary, Mohamed Ahmed Ali, Mohamed Ben Moussa, Mohamed Chenaoui, Mohamed El Sayes, Mohamed Elhadidi, Mohamed Gomaa Seadawy, Mohamed Hassan Abdoelraheem, Mohamed Hassany, Mohamed Houmed Aboubaker, Mohamed K.S., Mohamed Kamal, Mohamed Rhajaoui, Mohamed Seadawy, Mohamed Shamel, Mohamed Shemis, Mohammed K.S., Mohammed Walid Chemao Elfihri, Mohcine Bennani Mechita, Mokhtar Gomaa, Molalegne Bitew, Momoh M., Mona O.A. Alkarim, Monemo Pacome, Monilade Akinola, Monte A., Monuir G., Monze M., Mooko M., Morales A.N., Moreira-Soto Andres, Moriba Povogui, Mosepele Mosepele, Moses Chilufya, Moses Joloba, Moses Luutu, Mostafa Elouennass, Mostfa Elhoseiny, Mostfa Elnakib, Mostfa Yakout, Mouhcine Gardoul, Mouhssine Hemlali, Mouity Matoumba A., Mouna Ben Sassi, Mouna Safer, Mouneem Essabbar, Moustapha Mbow, Moustapha Nzamba Maloum, Moustapha Sakho, Moyinoluwa Odugbemi, Mtshali P., Mubemba B., Muchaneta Mugabe, Mufinda M., Muhammad Faisal, Muinah Adenike Fowora, Mukantwari Enatha, Muleya W., Muntaser elTayeb Ibrahim, Mupeta F., Murebwayire Clarisse, Mushal Ali, Mushal Allam, Mustapha Mouallif, Muuo S.N., Mwangomba W, Myriam Seffar, Mzumara T. E., N’dilimabaka N., Nabila Soara, Nabli A., Nadia El Mrimar, Nadia Rodrigues, Nadia Sitoe, Nafisatou Leye, Naguib A., Nalubamba K. S., Nancy M. El Guindy, Nandi Siegfried, Nardjes Hihi, Narjis Amar, Naryshkina E.A., Nathalia Endjala, Nathalia Garus-Oas, Nathan Kapata, Ndack Ndiaye, Ndahafa Frans, Ndam N.T., Ndéye Coumba Touré Kane, Ndiaye Ndack, Ndine Fainguem, Ndodo Nnaemeka, Ndumbu Pentikainen, Ndwiga L., Nedio Mabunda, Neff N., Negash A.A., Nejla Stambouli, Neto Z., Ngonga Dikongo A.M., Ngosa W., Ngozi Mirabel Otuonye, Niatou-Singa F. S., Nicaise T. Ndam, Nicholas Feasey, Nicholas Mwikwabe, Nicod J., Nicole Vidal, Nikki Freed, Nischay Mishra, Nissaf Ben Alaya, Nitin Savaliya, Noah Baker, Noé Patrick Mbondoukwe, Nokukhanya Mdlalose, Nonso Nduka, Noura M Abo Shama, Nourlil Jalal, Nsubuga Gideon, Ntuli N., Nuro Abilio, Nyam Itse Yusuf, Oby Wayoro, Ochwoto M., Ofonime Ebong, Ofori-Boadu L., Okomo Assoumou Marie Claire, Ola Elroby, Olabisi, Olabisi Ojo, Olajumoke Popoola, Olfert L., Olin Silander, Olufemi Obafemi, Olufemi Samuel Amoo, Olukunke Oluwasemowo, Olusola Anuoluwapo Akanbi, Oluwakemi Laguda-Akingba, Oluwatimilehin Adewumi, Omar Askander, Omar Elahmer-Abdulla Bashein-Rahma Algheriani-Ahlam Alarif, Omar S., Omilabu S., Omnia Kutkat, Omoare Adesuyi, Omondi Francis Carey, Onalethata Lesetedi, Ongera E., Ontlametse T. Bareng, Onwuamah C.K., Ope-Ewe O., Osama Mansour., Oscar Kanjerwa, Oteng F., Otmane Touzani, Oumaima Ait Si Mohammed, Oumy Diop, Ouna Ouadghiri, Ousseynou Gueye, Owusu-Nyantakyi C., Oyefolu A., Oyeronke Ayansola, P Nthiga, Palomba S., Panja L., Papa Alassane Diaw, Park D., Pasacaline Manga, Patel H., Patience Motshosi, Patoo M., Patrick Amoth, Patrick Descheemaeker, Patrick Mavingui, Patrick Tuyisenge, Pattoo M., Paul Dobi, Paul Liberator, Paulin N. Essone, Paulina Joãozinho da Costa Jarra Manneh, Pauline Yacine Sene, Paulo A Carralero RR Paixão J. P., Pavitra Roychoudhury, Peace O. Uche, Pei Zhou, Penda Malhado Diallo, Pereira A., Petas Akogbeto, Peter Bauer, Peter T. Skidmore, Petra Raimond, Phasha-Muchemenye Mmatshepho, Philip Ashton, Philip C., Philip El-Duah, Philip M. Soglo, Philip Wonder Phiri, Philipp Wagner, Philippe Colson, Philippe Dussart, Philippe Lavrard Meyer, Phillip Ashton, Pierre-Edouard Fournier, Piet Maes, Popova A. Yu., Portia Manangazira, Praise Adewumi, Qi Yang, Quaneeta Mohktar, Quansah E.B., Quashie P., Quedraogo, Rabia Maqsood, Rachel Githii, Rachid Abi, Rachid Benhida, Rachid El Jaoudi, Rachid Mentag, Rahaman A. Ahmed, Raiva Simbi, Rajiha Abubeker, Ramalia Chabi Nari, Ramon Lorenzo-Redondo, Ramy Galal, Raouf A., Raoul Saizonou, Raphael Lumembe, Ravena Mubichi, Regina Z. Cer, Reham Dawood, Reham Kassab, Rehema Liyai, Rehn A., Rei José Pereira, Reina Sikkema, Rfaki Abderrazak, Riad Mounir Armanious, Riadh Daghfous, Riadh Gouider, Richard Adegbola, Richard Lino Loro Lako, Richard Molenkamp, Richard Webby, Richmond Yeboah, Rick S., Rida Tagajdid, Rine Zeh Nfor, Rivalyn Nakoune Yandoko, Robert Rutayisire, Rodney S. Daniels, Rodrigue Bikangui, Rodrigue K. Kohoun, Rodrigue Kamga, Rodrigue Mintsa Nguema, Roger Shapiro, Rogers J., Rogers Kamulegeya, Rokaia Laamrti, Roméo Aimé Laclong Lontchi, Ronald Kiiza, Rosella De Nittis, Roshdy W.H., Rotimi Myrabelle Avome Houechenou, Roua Ben Othman, Rui Inndi, Saad M.A., Saaïd Amzazi, Sabin Nsanzimana, Sada Diallo, Sadji Y.A., Safae El Mazouri, Safae Elkochri, Safae Ghoulame, Safiatou Karidioula, Safietou Sankhe, Sahr Gevao, Sahr P.F., Saibu J.O., Saïdou Ouedraogo, Saiid S., Sainabou Laye Ndure, Sakoba Keita, Salah D., Salah Eldin Hussein, Salah H., Saleh A.A., Saleh M., Salifou Sourakatou, Sally Roberts, Salma Abid, Salma Sayed, Salou M., Salu O.B., Sam Lissauer, Sam O’neilla Oye Bingono, Samba Ndiour, Sameira M. Fageer, Samia Abdou Girgis, Samir M., Samir O., Samira Benkeroum, Samira F. Ibrahim, Samira M. Fageer, Samira Zoa Assoumou, Samirah Saiid, Samoel Ashimosi Khamadi, Samson Konongoi Limbaso, Samuel Armoo, Samuel Kirimunda, Samuel Sorie, Samwel Lifumo Symekher, Samwel Owaka, Sana Ferjani, Sanaa Alaoui-Amine, Sander Anna-Lena, Sankhe Safietou, Santiago Jiménez-Serrano, Santigie Kamara, Sara Chammam, Sara Mahmoud, Sarah Jefferies, Sarah Rubin, Sarah Stanley, Saraswathi Sathees, Saro Abdella, Sarra Chamman, Savannah Mwesigwa, Sawa H., Seadawy M.G., Sean Ellis, Sébastien Bontems, Sefetogi Ramaologa, Sekesai Zinyowera, Selassie Kumordjie, Sen Claudine Henriette Ngomtcho, Sena Awunyo, Serge Alain Sadeuh-Mba, Serigne Saliou Niane, Seth Okeyo, Settimia Altamura, Seyram Bless Agbenyo, Shah Mohamed Bakhash, Shahin Lockman, Shahinaz.A. Bedri, Shaimaa Soliman, Shalaby L., Shannon Wilson, Sharmini Muttaiyah, Sharon Abimbola, Sharon Hsu, Shcherbakova S.A., Shebbar Osiany, Shereen Shawky, Sherine Helmy, Shevtsova A.P., Shimaa moustafa, Shirlee Wohl, Shirley Johane, Sidonie A.M.Kagnissode, Silvanos Mukunzi Opanda, Sim Mayaphi, Simão Tchuda Bióté, Simeone Dal Monego, Simon Peter Ruhweza, Simone Eckstein, Sindayiheba Reuben, Sinyange N., Sivaramakrishna Rachakonda, Soa Fy Andriam, Sofia Viegas, Sogodogo, Sokhna Ndongo, Sola Ajibaye, Solomon Langat, Sombo Fwoloshi, Somda Soro Georgina Charlene, Sonal P. Henson, Sondes Haddad, Sonia V. Bedié, Sosedova E.A, Souad Kartti, Souissi Amira, Souleymane Mboup, Soundélé Maïté, Sounkalo Dao, Sourakatou Salifou, Srinivas Reddy Pallerla, Stefan Niemann, Steffen Borrmann, Stephen M. Eggan, Stephen Ochola, Subomi Olorunnimbe, Sudhir Bunga, Sujeewon C., Sunday Babatunde, Susan Engelbrecht, Susan Morpeth, Susan Taylor, Susann Handrick, Swaibu Gatare, Sylvie Melingui, Symeker S.L., Syntyche Devatchagni, Taha A.G., Taha Chouati, Taha Maatoug, Tahar Bajjou, Takada A., Taloa K.A., Tamrayehu Seyoum, Tan M., Tania Stander, Tanja Niemann, Tarek Aanniz, Tarek Refaat Elnagdy Raafat Zaher, Tatenda Takawira, Tato C., Tefera D.A., Tei-Maya Fred, Tesfaye Gelanew, Tesfaye Rufael, Thanh Le Viet, Thela Tefelo, Thérèse Kagoné, Thibaut Armel Cherif Gnimadi, Thiongo K., Thomas Briese, Thomas Van L., Thongbotho Mphoyakgosi, Toy Nwako, Thushan I de Silva, Tim Roloff, Timothy Blackmore, Tiziana Rollo, Tom Lutalo, Tomade A.M. Ibrahim, Tombolomako T. B., Tomiwa Adepetun, Tomkins-Tinch C., Tony Wawina-Bokalanga, Tope Sobajo, Touil Nadia, Touria Essayagh, Traoré, Triki H., Tsiry R., Tsiry Randriambolamanantsoa, Tumisang Madisa, Turki M., Ugwu C.A., Uyi Emokpae, Valeria Delli Carri, Valeria Micheli, Van Rooyen G., Vanaerschot M., Vanessa Magnussen, Vanessa Mohr, Vani Sathyendran, Veronica Playle, Viana R., Vickos U., Victor Max Corman, Victor Mukonka, Victor Ofula, Vida Ahyong, Vincent Appiah, Vincent Bours, Violette V. M’cormack, Virginia Hope, Vivi Hue-Trang Lieu, Vololoniaina Raharinosy, Wadegu Meshack, Wadonda N., Wadula J, Wael Ali, Waidi Sule, Wallace D Bulimo, Warren Cyrus Yiaba, Waruhiu C.N., Wasfi Fares, Webby R., Weldemariam A.G., Wendy Karen Jo, Woelfel R., Wolh S., Wurie I., Xiaoyun Ren, Xiuhua Wang, Ya.M. Krasnov, Yacine Amet Dia, Yacine DIA Seni Ndiaye, Yacob Mohamed Yusuf, Yacouba Sawadogo, Yadouleton, Yahya Maidane, Yakob Gebregziabher Tsegay, Yao Layibo, Yasser El Hady, Yassine Sekhsokh, Yassmin Moatasim, Yawo A. Sadji, Yeboah C., Youbi Mohammed, Yousif Rabih Makki, Youssef Akhoud, Yuri Ushijima, Yusuf Jimoh, Yvette Badou, Zablon J. Matoke D., Zablon J.O., Zaineb Hamzaoui, Zakia Regragui, Zara Wuduri, Zein Souma, Zeinab Ali Waberi, Zeinab S. Imam, Zekiba Tarnagda, Zemmouri Faouzia, Zhang J., Zhenghui Li, Zimmerman Maiga, Zohour Kasmy, Zong Minko O., Zorgani A., Zouheir Yassine, Zoukaneirii Issa, Zulu P., Zuzheng Xiang

